# Association of COVID-19 with neurodegenerative and neurological disorders in vaccinated and unvaccinated people

**DOI:** 10.64898/2026.10.30.26366919

**Authors:** Harry T. Mason, Mengxuan Zou, Emily Eyles, Jose Ignacio Cuitun Coronado, Paul Madley-Dowd, Marwa Al Arab, Yinghui Wei, Genevieve Cezard, Tom M. Palmer, Ameilia Green, Alex Walker, Simon Davy, Louis Fisher, Amir Mehrkar, Seb Bacon, Ben Goldacre, Nishi Chaturvedi, Jonathan A.C. Sterne, Rachel Denholm, Elizabeth Coulthard, Venexia Walker

**Affiliations:** Department of Population Health Sciences, Bristol Medical School, University of Bristol, Bristol, UK; The National Institute for Health and Care Research Applied Research Collaboration South West (NIHR ARC South West), UK; Academic Unit for Ageing and Stroke Research, University of Leeds, Bradford Teaching Hospitals NHS Foundation Trust, Bradford, UK; NIHR Bristol Biomedical Research Centre, Bristol, UK; MRC Integrative Epidemiology Unit, Bristol, UK; University of Exeter Medical School, Faculty of Health and Life Sciences, University of Exeter, Exeter, UK; NIHR Exeter Biomedical Research Centre, Exeter, UK; British Heart Foundation Cardiovascular Epidemiology Unit, Department of Public Health and Primary Care, University of Cambridge, Cambridge, UK; Victor Phillip Dahdaleh Heart and Lung Research Institute, University of Cambridge, Cambridge, UK; HDR UK Big Data for Complex Disease Data for Complex Disease driver programme, UK; The Bennett Institute for Applied Data Science, Nuffield Department of Primary Care Health Sciences, University of Oxford, Oxford, UK; MRC Unit for Lifelong Health and Ageing, University College London, London, UK; Health Data Research UK South-West, Bristol, UK; North Bristol NHS Trust, Bristol, UK; Department of Surgery, University of Pennsylvania Perelman School of Medicine, Philadelphia, Pennsylvania, USA

## Abstract

**Introduction:** COVID-19 has been linked to neuropathology, neurological symptoms, and diagnoses of neurological illnesses. Evidence regarding the long-term impact of COVID-19 on neurodegenerative and neurological disorders is still limited.

**Methods:** We investigated associations of COVID-19 with neurodegenerative and neurological disorders by vaccination status in both hospitalised patients and the general population in the largest and longest study to date. We used OpenSAFELY-TPP to access linked electronic health records from 18.3 million adults registered with English GPs. We defined three cohorts with up to 5.5 years follow-up: pre-vaccination (exposed to COVID-19 January 2020-June 2021), and vaccinated and unvaccinated (exposed June-December 2021). We estimated adjusted hazard ratios (aHRs) comparing the incidence of cognitive impairment and dementia after diagnosis of COVID-19 with before or without COVID-19. We also studied Parkinson’s disease, restless leg syndrome, rapid eye movement sleep disorder, motor neuron disease, multiple sclerosis, and migraine.

**Results:** Incidence of most outcomes was elevated during weeks 0-4 after COVID-19 (cognitive impairment/dementia aHRs; pre-vaccination: 4.02 (3.74-4.32)/5.05 (4.63-5.52), vaccinated: 2.24 (2.05-2.45)/2.05 (1.77-2.37), unvaccinated: 4.38 (3.45-5.55)/8.69 (6.13-12.32)). Incidence remained elevated up to 4 years after COVID-19, especially in those hospitalised with COVID-19 (cognitive impairment/dementia aHRs; pre-vaccination: 1.49 (1.35-1.64)/ 1.39 (1.21-1.60), vaccinated: 1.27 (1.04-1.54)/1.32 (1.02-1.71), unvaccinated: 1.55 (1.11-2.16)/1.48 (1.00-2.20)).

**Conclusion:** Incidence of cognitive impairment and dementia were elevated shortly after COVID-19. Incidence of cognitive impairment remained elevated in the longer term, particularly among unvaccinated individuals and/or those hospitalised with COVID-19. These patients may require additional assessment during routine care to evaluate persistent or new cognitive concerns.

## INTRODUCTION

At least 778 million people worldwide have been diagnosed with COVID-19, the disease caused by severe acute respiratory syndrome coronavirus 2 (SARS-CoV-2), including 25.1 million recorded cases in the UK ^1^. COVID-19 is associated with exacerbation of existing neurodegenerative disorders, as well as onset of new conditions ^2–11^. COVID-19 has also been linked to neuropathological changes, which can lead to neurodegenerative dementia^12^. Cognitive impairment is a common symptom of COVID-19^13^ and commonly reported among post-COVID sequele^14^, as well as a potential precursor to dementia^15^.

An increased risk of both cognitive impairment ^16,17^ and dementia ^12,18–22^ for up to 2 years after COVID-19 has been reported, despite dementia diagnoses commonly taking several months ^6,23^. In 2021, the average time for a dementia diagnosis in the UK after referral was reported to be 17.7 weeks^24^. We seek to expand upon these earlier studies by incorporating vaccination status and COVID-19 severity, an underexplored area for neurodegenerative and neurological disorders that has previously yielded useful insights for cardiovascular disease^25^, diabetes^26^ and mental health^27^. This has important implications for public health and dementia care provision^28^.

Using electronic health records for more than 18 million adults, we examined the associations between diagnosed COVID-19 with subsequent neurodegenerative and neurological disorders in three distinct cohorts defined by vaccination status and the COVID-19 variant they were likely exposed to. For cognitive impairment and dementia, we also examined associations by COVID-19 severity (defined by hospitalisation with COVID-19), as well as in subgroups for age, ethnicity, sex, prior history of COVID-19, previous cognitive impairment, high vascular risk, and previous Parkinson’s disease.

## METHODS

### Study design and data sources

We used OpenSAFELY-TPP^29–31^, which provides secure, privacy-protecting access to linked data from 36.7 million people registered with English general practices (GPs) using TPP SystmOne software, as part of the NHS England OpenSAFELY Analytics Service. These data include primary care data linked via pseudonymised NHS number to Secondary Uses Service (SUS) secondary care data, Office of National Statistics (ONS) Death Registry^32^, Second Generation Surveillance System (SGSS) COVID-19 testing data, and the Index of Multiple Deprivation 2019 (IMD). COVID-19 vaccination records (National Immunisation Management System) are available within TPP primary care data. No data from patients who registered a Type-1 Opt Out (https://docs.opensafely.org/type-one-opt-outs/) with their GP surgery were included in this study. More information on the patients available for selection in the Service is available in the [OpenSAFELY data](https://docs.opensafely.org/opensafely-data/) page of the documentation. In March 2020 the Secretary of State for Health and Social Care used powers under the UK Health Service (Control of Patient Information) Regulations 2002 to require organisations to process confidential patient information for the purposes of protecting public health, providing healthcare services to the public, and monitoring and managing the COVID-19 outbreak and incidents of exposure; this sets aside the requirement for patient consent. Ethics approval for this study was obtained from the Health Research Authority (REC reference 22/PR/0095) and the University of Bristol’s Faculty of Health Sciences Ethics Committee [reference 117269].

The primary outcomes were cognitive impairment and dementia (a composite outcome consisting of Alzheimer’s disease, vascular dementia, Lewy body dementia, other dementias and unspecified dementias). Other outcomes included dementia subtypes (Alzheimer’s disease, vascular dementia, Lewy body dementia), Parkinson’s disease, restless leg syndrome (also known as Willis-Ekbom disease), rapid eye movement (REM) sleep disorder, motor neurone disease (MND), multiple sclerosis, and migraine. Restless leg syndrome and REM sleep disorder were included as established risk conditions for Parkinson’s disease^33,34^. We examined multiple sclerosis and migraine to provide a broader assessment of neurological sequelae following COVID-19. Multiple sclerosis is a neuroinflammatory disease with a long-term degenerative component; while migraine, like cognitive impairment, is a common neurological condition without a degenerative component that have been linked to post-COVID sequelae^35^. Outcomes were defined using the earliest of: a SNOMED CT code (a structured clinical vocabulary to record clinical events) in primary care; start of a hospital admission with an ICD-10 code in any position, or death with an ICD-10 code in any position. Exceptions were Lewy body dementia, cognitive impairment and restless leg syndrome, which were defined using primary care data only as they are not well captured by four-character ICD-10 coding.

Date of COVID-19 was defined as the first of: confirmed diagnosis recorded in primary care; positive SARS-CoV-2 PCR or antigen test recorded in SGSS; start of a hospital admission with a confirmed diagnosis in any position, or death with SARS-CoV-2 infection listed as the primary or underlying cause. People with a hospital admission record including a confirmed diagnosis in the primary position within 28 days of first COVID-19 were defined as having had ‘hospitalised COVID-19’. All other COVID-19 diagnoses were defined as ‘non-hospitalised’.

Covariates identified as potential confounders included age, sex, ethnicity, deprivation (using IMD), smoking status, care home residence, obesity, health care work, GP practice visits in 2019, and binary indicators for comorbidities and prior history of relevant neurodegenerative and neurological conditions (**Supplementary Table 1**).

**Table 1:** Patient characteristics by cohort.

| Characteristics |  | Pre-vaccination cohort |  | Vaccinated cohort |  | Unvaccinated cohort |  |
|---|---|---|---|---|---|---|---|
|  |  | N (%) | COVID-19 diagnoses | N (%) | COVID-19 diagnoses | N (%) | COVID-19 diagnoses |
| Total, No. |  | 18,334,845 | 954,945 | 13,805,541 | 850,581 | 3,185,973 | 144,273 |
| Sex | Female | 9,175,491 (50.0%) | 510,669 | 7,172,259 (52.0%) | 472,365 | 1,325,469 (41.6%) | 75,735 |
|  | Male | 9,159,351 (50.0%) | 444,279 | 6,633,285 (48.0%) | 378,213 | 1,860,501 (58.4%) | 68,541 |
| Age, years | 18-49 | 9,424,977 (51.4%) | 598,569 | 5,852,553 (42.4%) | 471,327 | 2,531,487 (79.5%) | 120,753 |
|  | 50-64 | 4,543,317 (24.8%) | 234,531 | 3,885,861 (28.1%) | 264,261 | 469,623 (14.7%) | 19,011 |
|  | 65-84 | 3,782,811 (20.6%) | 97,071 | 3,539,961 (25.6%) | 104,313 | 165,087 (5.2%) | 3,933 |
|  | 85-110 | 583,731 (3.2%) | 24,777 | 527,169 (3.8%) | 10,683 | 19,779 (0.6%) | 579 |
| Ethnicity | Black* | 391,413 (2.1%) | 25,203 | 212,901 (1.5%) | 9,207 | 169,125 (5.3%) | 7,197 |
|  | Asian** | 1,209,615 (6.6%) | 116,175 | 800,067 (5.8%) | 41,709 | 333,081 (10.5%) | 9,633 |
|  | White | 14,892,327 (81.2%) | 742,473 | 11,765,103 (85.2%) | 747,555 | 2,020,503 (63.4%) | 113,415 |
|  | Chinese or Other | 359,811 (2.0%) | 15,183 | 199,881 (1.4%) | 8,571 | 166,767 (5.2%) | 3,201 |
|  | Mixed | 218,259 (1.2%) | 13,605 | 129,279 (0.9%) | 7,497 | 80,979 (2.5%) | 3,813 |
|  | Missing | 1,263,417 (6.9%) | 42,309 | 698,313 (5.1%) | 36,033 | 415,515 (13.0%) | 7,017 |
| IMD quintile | 1: most deprived | 3,566,814 (19.5%) | 236,730 | 2,271,324 (16.5%) | 134,940 | 956,298 (30.0%) | 45,042 |
|  | 2 | 3,622,134 (19.8%) | 205,488 | 2,571,108 (18.6%) | 155,544 | 761,016 (23.9%) | 34,050 |
|  | 3 | 3,969,708 (21.7%) | 190,326 | 3,059,454 (22.2%) | 183,138 | 638,466 (20.0%) | 28,320 |
|  | 4 | 3,726,162 (20.3%) | 171,858 | 3,002,886 (21.8%) | 187,422 | 484,302 (15.2%) | 21,438 |
|  | 5: least deprived | 3,450,018 (18.8%) | 150,534 | 2,900,760 (21.0%) | 189,540 | 345,894 (10.9%) | 15,426 |
| Smoking | Never | 8,405,445 (45.8%) | 475,953 | 6,422,469 (46.5%) | 414,783 | 1,321,557 (41.5%) | 57,147 |
|  | Formerly | 6,006,015 (32.8%) | 310,935 | 5,008,869 (36.3%) | 327,081 | 623,643 (19.6%) | 41,961 |
|  | Currently | 3,152,193 (17.2%) | 128,997 | 1,945,455 (14.1%) | 92,541 | 867,003 (27.2%) | 36,825 |
|  | Missing | 771,189 (4.2%) | 39,063 | 428,745 (3.1%) | 16,173 | 373,773 (11.7%) | 8,337 |
| Region | East | 4,178,943 (22.8%) | 210,351 | 3181,857 (23%) | 176,289 | 710,775 (22.3%) | 32,181 |
|  | East Midlands | 3,192,759 (17.4%) | 180,207 | 2,422,839 (17.5%) | 157,473 | 514,203 (16.1%) | 27,609 |
|  | London | 1,173,231 (6.4%) | 61,329 | 684,537 (5.0%) | 34,611 | 435,549 (13.7%) | 10,083 |
|  | North East | 909,993 (5.0%) | 59,697 | 679,101 (4.9%) | 50,169 | 141,729 (4.4%) | 7,389 |
|  | North West | 1714,089 (9.3%) | 109,947 | 1,312,923 (9.5%) | 91,221 | 234,723 (7.4%) | 12,459 |
|  | South East | 1231,485 (6.7%) | 45,813 | 956,415 (6.9%) | 56,097 | 192,495 (6.0%) | 8,313 |
|  | South West | 2,575,833 (14.0%) | 74,043 | 2,146,629 (15.5%) | 129,033 | 320,241 (10.1%) | 16,743 |
|  | West Midlands | 741,663 (4.0%) | 52,929 | 486,657 (3.5%) | 29,019 | 177,813 (5.6%) | 7,917 |
|  | Yorkshire/Humber | 2,616,849 (14.3%) | 160,623 | 1,934,583 (14.0%) | 126,657 | 458,433 (14.4%) | 21,579 |
| GP practice visits in 2019 | 0 | 5,544,039 (30.2%) | 228,441 | 3,704,547 (26.8%) | 214,995 | 1,787,439 (56.1%) | 47,829 |
|  | 1-5 | 7,104,111 (38.7%) | 403,365 | 5,561,769 (40.3%) | 363,213 | 929,049 (29.2%) | 59,469 |
|  | 6+ | 5,686,695 (31.0%) | 323,139 | 4,539,225 (32.9%) | 272,367 | 469,485 (14.7%) | 36,975 |
|  | Care home resident | 88,041 (0.5%) | 13,959 | 56,565 (0.4%) | 2,697 | 2,895 (0.1%) | 99 |
|  | Healthcare worker | 555,411 (3.0%) | 60,441 | 472,377 (3.4%) | 37,575 | 19,551 (0.6%) | 1,839 |

IMD: Index of Multiple Deprivation 2019
\* or Black British
\*\* or Asian British

### Study population

Three cohorts were defined (**Supplementary Table 2**). The ‘pre-vaccination’ cohort was followed from January 1^st^ 2020 (baseline) until the earliest of: May 31^st^ 2025, date of outcome, date of deregistration and date of death. Exposure in the pre-vaccination cohort was defined as a COVID-19 diagnosis recorded between baseline and the earliest date of: eligibility for COVID-19 vaccination, date of first vaccination, and June 18^th^ 2021 (when all adults became eligible for vaccination). Follow-up in the ‘vaccinated’ cohort started at the later of June 1^st^ 2021 (baseline) and two weeks after a second COVID-19 vaccination and ended at the earliest of May 31^st^ 2025, date of outcome, date of deregistration and date of death. Follow-up in the ‘unvaccinated’ cohort started at the later of June 1^st^ 2021 (baseline) and 12 weeks after vaccination eligibility, and ended at the earliest of May 31^st^ 2025, date of outcome, date of deregistration, and date of death. Exposure in the vaccinated and unvaccinated cohorts was defined as a COVID-19 diagnosis recorded between the cohort baseline and December 14^th^ 2021 (when Omicron become the dominant variant).

**Table 2:** Neurodegenerative and neurological disorders following diagnosis of COVID-19 by cohort, overall, and by COVID-19 severity.

| Outcome | COVID-19 severity | Pre-vaccination cohort |  | Vaccinated cohort |  | Unvaccinated cohort |  |
|---|---|---|---|---|---|---|---|
|  |  | Event/ person-years | Incidence rate/ 100,000 person-years | Event/ person-years | Incidence rate/ 100,000 person-years | Event/ person-years | Incidence rate/ 100,000 person-years |
| **Cognitive impairment** | No COVID-19 | 432501/75839144 | 570 | 326361/42717163 | 764 | 18195/9569158 | 190 |
|  | Non-hospitalised COVID-19 | 14631/3262543 | 448 | 12699/2665436 | 476 | 1239/419036 | 296 |
|  | Hospitalised COVID-19 | 2439/131292 | 1858 | 855/29310 | 2917 | 225/21502 | 1046 |
| **Dementia** | No COVID-19 | 250755/38481639 | 652 | 184047/26173329 | 703 | 5133/2021449 | 254 |
|  | Non-hospitalised COVID-19 | 4347/1122022 | 387 | 3171/1209909 | 262 | 93/60719 | 153 |
|  | Hospitalised COVID-19 | 1575/98693 | 1596 | 573/25591 | 2239 | 99/10547 | 939 |
| **Alzheimer's disease** | No COVID-19 | 130473/38629682 | 338 | 100455/26263482 | 382 | 1989/2024417 | 98 |
|  | Non-hospitalised COVID-19 | 1869/1126306 | 166 | 1761/1211780 | 145 | 27/60790 | 44 |
|  | Hospitalised COVID-19 | 597/100413 | 595 | 237/26004 | 911 | 27/10626 | 254 |
| **Vascular dementia** | No COVID-19 | 46503/38770103 | 120 | 32103/26361146 | 122 | 939/2025823 | 46 |
|  | Non-hospitalised COVID-19 | 981/1128183 | 87 | 597/1213672 | 49 | 27/60818 | 44 |
|  | Hospitalised COVID-19 | 405/100879 | 401 | 117/26213 | 446 | 15/10652 | 141 |
| **Lewy body dementia** | No COVID-19 | 4881/38823696 | 13 | 3483/26393499 | 13 | 51/2026761 | 3 |
|  | Non-hospitalised COVID-19 | 69/1129682 | 6 | 87/1214320 | 7 | 3/60844 | 5 |
|  | Hospitalised COVID-19 | 27/101464 | 27 | 9/26350 | 34 | 0/10676 | 0 |
| **Parkinson's disease** | No COVID-19 | 37611/38605733 | 97 | 28269/26239378 | 108 | 621/2022733 | 31 |
|  | Non-hospitalised COVID-19 | 735/1125157 | 65 | 747/1210304 | 62 | 21/60762 | 35 |
|  | Hospitalised COVID-19 | 195/100330 | 194 | 57/26011 | 219 | 9/10622 | 85 |
| **Restless leg syndrome** | No COVID-19 | 32139/78069145 | 41 | 21171/44230075 | 48 | 1485/9696004 | 15 |
|  | Non-hospitalised COVID-19 | 1581/3345273 | 47 | 1365/2725258 | 50 | 141/425811 | 33 |
|  | Hospitalised COVID-19 | 129/144228 | 89 | 39/32842 | 119 | 15/22706 | 66 |
| **Rapid eye movement sleep disorder** | No COVID-19 | 132747/77952375 | 170 | 94047/44120953 | 213 | 6993/9682887 | 72 |
|  | Non-hospitalised COVID-19 | 6795/3331899 | 204 | 6327/2713096 | 233 | 525/424696 | 124 |
|  | Hospitalised COVID-19 | 1605/137387 | 1168 | 471/30890 | 1525 | 171/21977 | 778 |
| **Migraine** | No COVID-19 | 228801/70971846 | 322 | 132489/39954884 | 332 | 23295/9104664 | 256 |
|  | Non-hospitalised COVID-19 | 15501/2944042 | 527 | 10905/2388664 | 457 | 2121/377788 | 561 |
|  | Hospitalised COVID-19 | 489/129088 | 379 | 129/29334 | 440 | 93/19967 | 466 |
| **Motor neurone disease** | No COVID-19 | 4875/78662131 | 6 | 3345/44626363 | 7 | 147/9717797 | 2 |
|  | Non-hospitalised COVID-19 | 135/3367131 | 4 | 117/2745544 | 4 | 3/427301 | 1 |
|  | Hospitalised COVID-19 | 21/146390 | 14 | 9/33630 | 27 | 3/22880 | 13 |
| **Multiple sclerosis** | No COVID-19 | 11325/78405247 | 14 | 6825/44457379 | 15 | 951/9704335 | 10 |
|  | Non-hospitalised COVID-19 | 501/3358529 | 15 | 441/2735550 | 16 | 63/426500 | 15 |
|  | Hospitalised COVID-19 | 45/145253 | 31 | 15/33199 | 45 | 9/22776 | 40 |

People eligible for each cohort were registered with an English GP for at least six months before baseline and were alive with a known age between 18 and 110 years, sex, deprivation, and region at baseline. For the outcomes of dementia and Parkinson’s disease, we restricted it to people aged 50 to 110 years; otherwise, we included all people aged 18 to 110 years. People were excluded from each cohort if they had any record of COVID-19 before baseline and studied separately in a subgroup analysis. In the vaccinated cohort, people were excluded if they received a vaccination before the start of the vaccine rollout on December 8^th^ 2020 (indicating an error or participation in a randomised trial); their second dose was dated before their first dose vaccination (contradictory vaccine record); their second dose was less than three weeks after their first dose; or they received mixed vaccine brands before this was permitted on May 7^th^ 2021. In the unvaccinated cohort, people were excluded if they had any record of a COVID-19 vaccination before June 1^st^ 2021. Vaccine eligibility was defined using the Joint Committee on Vaccination and Immunisation (JCVI) groupings. People who could not be assigned to a JCVI group were excluded.

### Statistical analyses

For each cohort, baseline characteristics were described, and numbers of outcome events, person-years of follow-up and incidence rates (per 100,000 person-years) before and after all, hospitalised and non-hospitalised COVID-19 were tabulated. Time to first event was analysed for each outcome. Cox models were fitted with calendar time scale using the cohort-specific baseline as the origin. Hazard ratios (HRs) for follow-up after all, hospitalised, and non-hospitalised COVID-19, versus before or without COVID-19, were estimated, splitting follow-up into: 0-4 weeks (excluding day 0), 5-26 weeks, 27-52 weeks, 1-2 years, 2-3 years, 3-4 years after COVID-19 for all cohorts and additionally 4-5.5 years (282 weeks) after COVID-19 for the pre-vaccination cohort. If there were less than 12 events in any of the first three time periods (i.e. 0-4 weeks (excluding day 0), 5-26 weeks, 27-52 weeks), we used a single time period (i.e., 0-1 years (excluding day 0)) in place of the three time periods. For computational efficiency, we included all people with the outcome event, all people with the exposure (COVID-19 diagnosis), and a 20% random sample of people who were not diagnosed with COVID-19 and in whom the outcome event was not recorded. We used inverse probability weights to adjust for the sampling and derived confidence intervals using robust standard errors. For each outcome and cohort, we estimated: (i) age and sex adjusted; and (ii) maximally adjusted HRs including all covariates. Restricted cubic splines were used to account for age unless otherwise specified. All models were stratified by region to construct risk sets within-region, accounting for between-region variation in the baseline hazard. Models with less than 50 events overall were excluded.

Subgroup analyses according to age group, sex, ethnicity, and COVID-19 history were conducted for all outcomes. Additional outcome specific subgroups were also investigated: history of Parkinson’s risk conditions (defined as REM sleep disorder and/or restless leg syndrome) for the outcome Parkinson’s disease; history of cognitive impairment for the outcome dementia; history of Parkinson’s disease for the outcome dementia; and high vascular risk (defined as diabetes and/or hypertension) for the outcomes cognitive impairment, dementia, and the dementia subtypes. We calculated absolute excess risk a year after COVID-19 and weighted by the proportion of people in age and sex strata in the pre-vaccination cohort (**Supplementary Text 1**).

The study was conducted in OpenSAFELY v1.56.3, Python v3.10.12, R v4.4.3, and Stata/MP v16.1 according to a pre-specified protocol. Our protocol, analysis code, and code lists are available: https://github.com/opensafely/post-covid-neurodegenerative. Deviations from the protocol are listed in **Supplementary Text 2**. All outputs were subjected to OpenSAFELY disclosure controls, including rounding where appropriate: https://docs.opensafely.org/releasing-files.

## RESULTS

The pre-vaccination cohort included 18,334,845 people (9,175,491 million [50.0%] female, 9,159,351 million [50.0%] male, median [IQR] age 49 [34-64] years), 954,945 of whom received a COVID-19 diagnosis (**Table 1, Supplementary Figure 1**). The cohort included 391,413 (2.1%) Black or Black British individuals; 1,209,615 (6.6%) Asian or Asian British individuals; 14,892,327 (81.2%) White individuals; 218,259 (1.2%) mixed ethnicity individuals, 359,811 (2.0%) Chinese and other ethnic group individuals, and 1,263,417 (6.9%) individuals with no recorded ethnicity. The vaccinated cohort included 13,805,541 people (7,172,259 [52.0%] female, 6,633,285 [48.0%] male, median [IQR] age 53 [38-67] years), 850,581 of whom received a COVID-19 diagnosis (**Table 1**). The cohort included 212,901 (1.5%) Black or Black British individuals; 800,067 (5.8%) Asian or Asian British individuals, 11,765,103 (85.2%) White individuals, 129,279 (0.9%) mixed ethnicity individuals, 199,881 (1.4%) Chinese and other ethnic group individuals, and 698,313 (5.1%) individuals with no recorded ethnicity. The unvaccinated cohort included 3,185,973 million people (1,325,469 [41.6%] female, 1,860,501 [58.4%] male, median [IQR] age 36 [28-47] years), 144,273 of whom received a COVID-19 diagnosis (**Table 1**). The cohort included 169,125 (5.3%) Black or Black British individuals; 333,081 (10.5%) Asian or Asian British individuals, 2,020,503 (63.4%) White individuals, 80,979 (2.5%) mixed ethnicity individuals, 166,767 (5.2%) Chinese or other ethnic group individuals, and 415,515 (13.0%) individuals with no recorded ethnicity. **Supplementary Table 3** summarises participants’ medical history by cohort. Differences in demographic characteristics and medical history between the cohorts reflects factors associated with COVID-19 vaccine uptake, with the unvaccinated cohort generally being younger, more deprived and less likely to interact with health services.^36,37^

**Table 3:** Maximally adjusted hazard ratios and 95% confidence intervals for cognitive impairment and dementia following diagnosis of COVID-19 by cohort following: any diagnosis of COVID-19, diagnosis of hospitalised COVID-19, and diagnosis of non-hospitalised COVID-19.

| Outcome | Time since COVID-19 | Pre-vaccination cohort | Vaccinated cohort | Unvaccinated cohort |
|---|---|---|---|---|
| **All COVID-19** |  |  |  |  |
| **Cognitive impairment** | 0-4 weeks | 4.02 (3.74-4.32) | 2.24 (2.05-2.45) | 4.38 (3.45-5.55) |
|  | 5-26 weeks | 2.00 (1.92-2.09) | 1.40 (1.33-1.47) | 1.90 (1.62-2.22) |
|  | 27-52 weeks | 1.40 (1.33-1.46) | 1.03 (0.98-1.08) | 1.47 (1.26-1.72) |
|  | 1-2 years | 1.20 (1.16-1.24) | 0.96 (0.93-0.99) | 1.36 (1.21-1.52) |
|  | 2-3 years | 1.09 (1.06-1.13) | 1.05 (1.02-1.09) | 1.56 (1.41-1.71) |
|  | 3-4 years | 1.13 (1.09-1.17) | 1.15 (1.11-1.19) | 1.37 (1.22-1.55) |
|  | 4-5.5 years | 1.29 (1.23-1.34) | - | - |
| **Dementia** | 0-4 weeks | 5.05 (4.63-5.52) | 2.05 (1.77-2.37) | 8.69 (6.13-12.32) |
|  | 5-26 weeks | 2.00 (1.87-2.14) | 1.13 (1.04-1.24) | 1.44 (0.98-2.13) |
|  | 27-52 weeks | 1.44 (1.34-1.55) | 0.97 (0.89-1.06) | 1.42 (0.97-2.09) |
|  | 1-2 years | 1.18 (1.12-1.26) | 0.89 (0.84-0.95) | 1.14 (0.83-1.58) |
|  | 2-3 years | 1.12 (1.05-1.19) | 0.88 (0.83-0.94) | 1.00 (0.70-1.44) |
|  | 3-4 years | 1.13 (1.06-1.21) | 0.86 (0.79-0.93) | 1.40 (0.95-2.05) |
|  | 4-5.5 years | 1.06 (0.97-1.15) | - | - |
| **Hospitalised COVID-19** |  |  |  |  |
| **Cognitive impairment** | 0-4 weeks | 7.95 (6.97-9.07) | 4.60 (3.66-5.78) | 9.06 (5.95-13.82) |
|  | 5-26 weeks | 3.36 (3.04-3.72) | 2.58 (2.21-3.01) | 3.88 (2.84-5.30) |
|  | 27-52 weeks | 2.04 (1.82-2.30) | 2.00 (1.70-2.35) | 2.00 (1.37-2.92) |
|  | 1-2 years | 1.62 (1.48-1.78) | 1.33 (1.15-1.54) | 1.75 (1.30-2.34) |
|  | 2-3 years | 1.38 (1.25-1.52) | 1.41 (1.22-1.64) | 1.68 (1.27-2.21) |
|  | 3-4 years | 1.49 (1.35-1.64) | 1.27 (1.04-1.54) | 1.55 (1.11-2.16) |
|  | 4-5.5 years | 1.31 (1.15-1.49) | - | - |
| **Dementia** | 0-4 weeks | 11.78 (10.44-13.29) | 8.12 (6.58-10.02) | 3.46 (2.59-4.64) |
|  | 5-26 weeks | 2.90 (2.56-3.30) | 2.34 (1.93-2.83) |  |
|  | 27-52 weeks | 1.98 (1.71-2.28) | 2.06 (1.68-2.53) |  |
|  | 1-2 years | 1.69 (1.50-1.90) | 1.42 (1.18-1.70) | 1.27 (0.77-2.10) |
|  | 2-3 years | 1.38 (1.21-1.58) | 1.44 (1.18-1.75) | 1.48 (1.00-2.20) |
|  | 3-4 years | 1.39 (1.21-1.60) | 1.32 (1.02-1.71) |  |
|  | 4-5.5 years | 1.32 (1.10-1.58) | - | - |
| **Non-hospitalised COVID-19** |  |  |  |  |
| **Cognitive impairment** | 0-4 weeks | 3.31 (3.04-3.61) | 2.06 (1.87-2.26) | 3.54 (2.67-4.71) |
|  | 5-26 weeks | 1.83 (1.74-1.92) | 1.33 (1.27-1.40) | 1.61 (1.34-1.92) |
|  | 27-52 weeks | 1.32 (1.25-1.39) | 0.98 (0.93-1.04) | 1.40 (1.18-1.66) |
|  | 1-2 years | 1.15 (1.11-1.19) | 0.94 (0.91-0.98) | 1.31 (1.16-1.47) |
|  | 2-3 years | 1.06 (1.02-1.10) | 1.04 (1.01-1.08) | 1.54 (1.39-1.71) |
|  | 3-4 years | 1.10 (1.06-1.14) | 1.14 (1.10-1.19) | 1.35 (1.19-1.54) |
|  | 4-5.5 years | 1.28 (1.23-1.34) | - | - |
| **Dementia** | 0-4 weeks | 2.99 (2.63-3.41) | 1.16 (0.94-1.43) | 1.31 (0.92-1.87) |
|  | 5-26 weeks | 1.79 (1.66-1.94) | 1.00 (0.91-1.10) |  |
|  | 27-52 weeks | 1.31 (1.20-1.43) | 0.86 (0.78-0.95) |  |
|  | 1-2 years | 1.07 (1.00-1.15) | 0.85 (0.80-0.91) | 1.12 (0.74-1.69) |
|  | 2-3 years | 1.06 (0.99-1.14) | 0.85 (0.79-0.91) | 1.01 (0.71-1.44) |
|  | 3-4 years | 1.08 (1.00-1.16) | 0.83 (0.76-0.90) |  |
|  | 4-5.5 years | 1.00 (0.90-1.10) | - | - |

- Time period falls outside of cohort follow-up

**Figure 1:**
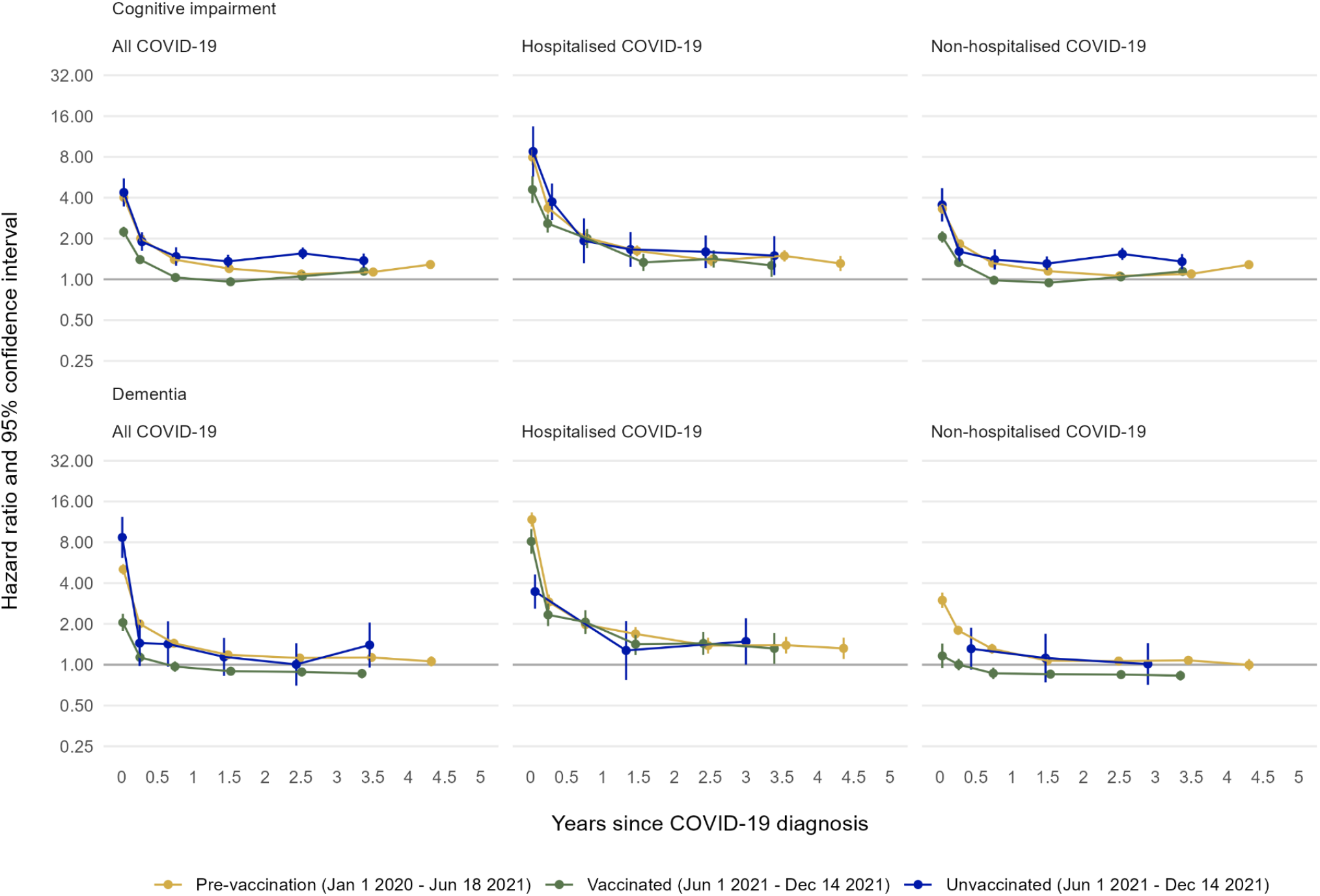
Maximally adjusted hazard ratios and 95% CIs for cognitive impairment and dementia following diagnosis of COVID-19 by cohort, overall, and by COVID-19 severity.

The incidences of both cognitive impairment and dementia were more elevated following hospitalised COVID-19 than before or without COVID-19 in all cohorts (**Table 2**). There were 449,571, 339,915, and 19,659 total diagnoses of cognitive impairment and 256,677, 187,791, and 5,325 diagnoses of dementia in the pre-vaccination, vaccinated, and unvaccinated cohorts respectively.

### Comparisons of event rates after diagnosis of COVID-19 vs before or without COVID-19

Maximally adjusted HRs (aHRs) comparing the incidence of each outcome after COVID-19 with the incidence before or without COVID-19 did not differ substantially from the age- and sex-adjusted HRs (**Supplementary Table 4**). For all figures, aHRs and 95% confidence intervals (95% CI) are plotted against the median time from date of COVID-19 to date of outcome in each cohort.

#### Cognitive Impairment

Incidence of cognitive impairment was elevated during weeks 0-4 after COVID-19 in the pre-vaccination (aHR: 4.02; 95% CI: 3.74-4.32) and unvaccinated (aHR: 4.38; 95% CI: 3.45-5.55) cohorts (**Table 3, Figure 1**). Incidence remained elevated up to 4 years after COVID-19 in the unvaccinated cohort (aHR: 1.37; 95% CI: 1.22-1.55) and up to 5.5 years in the pre-vaccination cohort (aHR 1.29; 95% CI 1.23-1.34). The aHR for cognitive impairment was lower in the vaccinated than the pre-vaccination and unvaccinated cohorts (aHRs 2.24 [95% CI 2.05-2.45] and 1.50 [1.33-1.47] during weeks 0-4 and 5-26 respectively). Subsequent aHRs in the vaccinated cohort were close to 1, although the aHR was 1.15 (95% CI 1.11-1.19) 3-4 years after COVID-19. The elevation in incidence of cognitive impairment was greatest amongst those hospitalised with COVID-19, which was the only subgroup in which all three cohorts had an elevated risk after a year (**Table 3, Figure 1**). The aHRs for the unvaccinated cohort were consistently greater than 1, even in those not hospitalised with COVID-19 (**Figure 1**).

#### Dementia

Incidence of dementia after diagnosed COVID-19 was substantially elevated during weeks 0-4 in all cohorts (pre-vaccination aHR: 5.05; 95% CI:4.63-5.52, vaccinated aHR: 2.05; 95% CI: 1.77-2.37, unvaccinated aHR: 8.69; 95% CI:6.13-12.3; **Table 3, Figure 1**). Incidence of dementia remained elevated up to 4 years after COVID-19 in the pre-vaccination cohort (aHR: 1.13; 95% CI:1.09-1.17) but was similar to before or without COVID-19 within a year in the vaccinated and unvaccinated cohorts. The aHRs were much more elevated amongst those hospitalised with COVID-19 and remained elevated throughout follow-up. For instance, the aHRs for dementia up to 4 years after COVID-19 were: 1.39 (95% CI:1.21-1.60), 1.32 (95% CI:1.02-1.71), and 1.48 (95% CI:1.00-2.20) in the pre-vaccination, vaccinated, and unvaccinated cohorts respectively (**Table 3, Figure 1**).

#### Subgroup analyses

Incidence of cognitive impairment and dementia after COVID-19 amongst people with a history of COVID-19, did not differ with that before or without COVID-19, with one exception: the aHR of cognitive impairment in the vaccinated cohort in the first 4 weeks after COVID-19 was 2.19 (95% CI: 1.39-3.44) (**Supplementary Figure 2, Supplementary Table 5**). Incidences of cognitive impairment and dementia were broadly similar across age (**Supplementary Figure 3, Supplementary Tables 6-7**), sex (**Supplementary Figure 4, Supplementary Table 8**), ethnicity (**Supplementary Figure 5, Supplementary Tables 9-10**) and vascular risk (**Supplementary Figure 6, Supplementary Tables 11**). Incidence of dementia was more elevated in people without a history of cognitive impairment compared with those with a history of cognitive impairment (**Supplementary Figure 7, Supplementary Tables 12**). Incidence of dementia was comparable in people with and without a history of Parkinson’s disease (**Supplementary Figure 8, Supplementary Table 13**).

**Figure 2:**
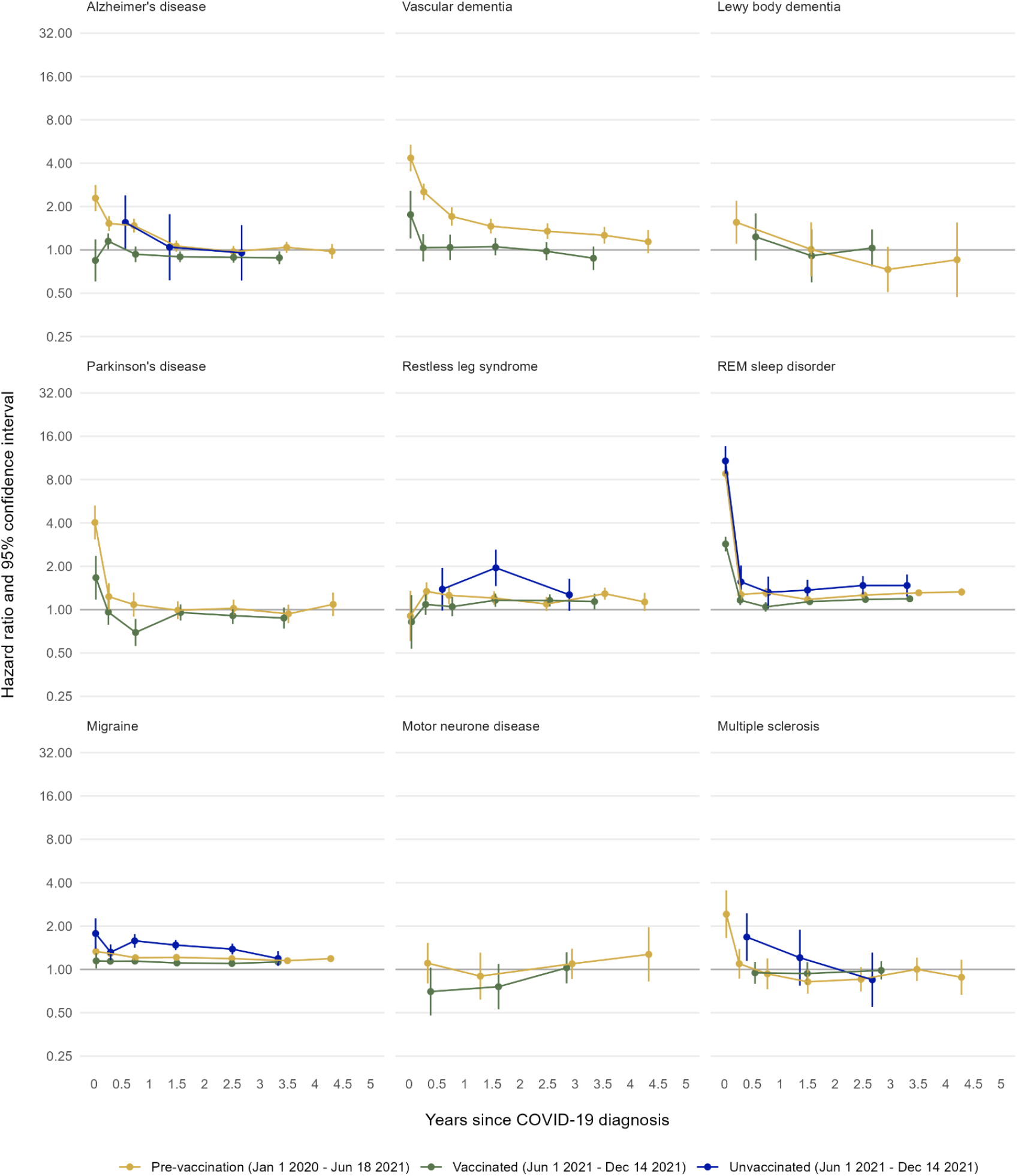
Maximally adjusted hazard ratios and 95% CIs for other neurodegenerative and neurological conditions following diagnosis of COVID-19 by cohort.

#### Other neurodegenerative and related risk conditions

The incidence of Alzheimer’s disease after COVID-19 compared with before or without COVID-19 was similar to the incidence of any dementia and most elevated in people with hospitalised COVID-19 (**Figure 2, Supplementary Figure 9, Supplementary Table 14**). However, the aHR of vascular dementia was markedly elevated in the pre-vaccination cohort, remaining elevated for up to 4 years (**Figure 2, Supplementary Table 14**). The incidence of Lewy body dementia after COVID-19 was similar to before or without COVID-19 (**Figure 2, Supplementary Figure 9, Supplementary Table 14**). Incidence of Parkinson’s disease was elevated for weeks 0-4 after COVID-19 and broadly comparable with before or without COVID-19 from week 4 onwards, while incidence of restless leg syndrome was broadly comparable with before or without COVID-19 throughout follow-up (**Figure 2, Supplementary Figure 10, Supplementary Table 15**). Incidence of Parkinson’s disease after COVID-19 in people with and without Parkinson’s risk conditions was similar (**Figure 2, Supplementary Figure 11, Supplementary Table 16**). The aHRs for REM sleep disorder were elevated after COVID-19, and remained consistently elevated from 4 weeks onwards (especially in the pre-vaccination and unvaccinated cohorts) (**Figure 2, Supplementary Figure 10, Supplementary Table 15**). Incidence of migraine had a small elevation throughout follow-up in the unvaccinated cohort but was similar to before or without COVID-19 for the other cohorts, with the exception of the first 4 weeks after hospitalised COVID-19, which was elevated in all cohorts (**Figure 2, Supplementary Figure 12, Supplementary Table 17)**. Incidence of motor neurone disease after COVID-19 was comparable with before or without COVID-19 throughout follow-up, while incidence of multiple sclerosis was elevated for weeks 0-4 after COVID-19 and then broadly comparable with before or without COVID-19 from week 4 onwards (**Figure 2, Supplementary Figure 12, Supplementary Table 17**).

### Absolute excess risk

Estimated excess risks of cognitive impairment a year after COVID-19, standardised to the age and sex distribution of the pre-vaccination cohort, were 493, 181, and 333 per 100,000 people in the pre-vaccination, vaccinated, and unvaccinated cohorts, respectively (**Supplementary Figure 13**). The equivalent estimated excess risks of dementia were 665, 83, and 453 people per 100,000. The excess risk increased with age and was higher for women than men for both outcomes.

## DISCUSSION

In this cohort study of more than 18 million people with up to 5.5 years of follow-up, incidence of cognitive impairment remained elevated up to 5.5 years after COVID-19 in the pre-vaccination cohort and up to 4 years after COVID-19 in the unvaccinated cohort. While incidence of cognitive impairment was initially elevated up to 4 weeks after COVID-19 in the vaccinated cohort, after the first year, incidence was comparable to before or without COVID-19. Incidence of cognitive impairment was more elevated after hospitalised COVID-19, compared with non-hospitalised COVID-19, in both the pre-vaccination and vaccinated cohorts. However, incidence of cognitive impairment was comparable after hospitalised and non-hospitalised COVID-19 in the unvaccinated cohort. Incidence of dementia remained elevated up to 4 years after COVID-19 in the pre-vaccination cohort but was similar to before or without COVID-19 within a year after COVID-19 in the vaccinated and unvaccinated cohorts. Incidence of dementia was more elevated after hospitalised COVID-19 for all cohorts.

Incidence of Alzheimer’s disease, Parkinson’s disease, and multiple sclerosis were all elevated initially after COVID-19 but were broadly comparable to before or without COVID-19 beyond the initial elevation; while incidence of Lewy Body dementia, restless leg syndrome and motor neurone disease were comparable to before or without COVID-19 throughout follow-up. Incidence of vascular dementia was elevated for up to 4 years in the pre-vaccinated cohort, incidence of migraine was elevated for up to 2.5 years in the unvaccinated cohort, and incidence of REM sleep disorder was elevated throughout follow-up in all cohorts.

Post-COVID sequelae can affect nearly every organ system^38^ and are defined as any new or continuing symptom persisting beyond 12 weeks^39^. While this study does not directly evaluate post-COVID sequelae, cognitive impairment is a common component of post-COVID sequelae definitions^40^, and cognitive scores have previously shown to worsen at a greater rate amongst those hospitalised with COVID-19^41^. Here, we show that incidence of cognitive impairment is elevated in the first year after COVID-19, including within the first 12 weeks. Vaccines have previously been shown to reduce the risk of post-COVID sequelae^42,43^, and specifically to help with cognitive impairment in mice^44^. Here we show the importance of vaccination specifically in cognitive impairment after COVID-19 in humans, in the longest study to address this question to date and identify important differences by COVID-19 severity. Several existing studies have investigated the association of COVID-19 and dementia^5,18,20,45–58^, including dementia subtypes (i.e. Alzheimer’s disease^5,18,45–53^, vascular dementia^59^, and dementia with Lewy bodies^54^). Taquet et al.^6^ estimated an aHR of 2.33 (95% CI: 1.77-3.07) for dementia up to six months after COVID-19, while Zhang et al.^60^ estimated an aHR of 1.65 (95% CI: 1.39–1.95) for dementia up to two years after COVID-19 in a meta-analysis across a comparable sample size (26.4 million). Shan et al.^61^ estimated an aHR of 2.42 (95% CI: 1.15-5.10) for vascular dementia amongst those hospitalised with COVID-19 with a median follow up of 24 months, finding vascular dementia to be the most affected dementia subtype. Our findings from the pre-vaccination cohort, which best reflect the people in the previous studies (exposed early in the pandemic, before vaccinations were available), are broadly consistent with these previous reports and provide additional follow-up up to 5.5 years. Other neurodegenerative diseases and related risk conditions have been less studied due to the limited sample sizes for these outcomes. Nevertheless, associations have previously been reported between COVID-19 and Parkinson’s disease^5,45,49,62–65^, sleep disturbances^66–69^, multiple sclerosis^64,70–72^, and motor neuron disease^73,74^. Among our other outcomes, we observed an elevation in the incidence of migraine and REM sleep disorder following COVID-19. Like cognitive impairment, migraine can manifest as a neurological symptom during COVID-19^69^, and can also be associated with post-COVID sequelae^75^.

A key strength of this study is the large sample (>18 million people) and detailed electronic health record data, combined with a long follow-up time, which enabled comparisons by vaccination status and time period after COVID-19 diagnosis in several relevant subgroups. Linkage of primary and secondary care data with national SARS-CoV-2 testing data improved ascertainment of COVID-19 outcomes and covariates, and enabled us to study people who were and were not hospitalised with their COVID-19. However, despite the wealth of data that we had access to, incidence rates of many neurodegenerative outcomes are low meaning many of our analyses were underpowered. This limitation is particularly noticeable in the unvaccinated cohort, where around 80% of the cohort were under 50 years old. Furthermore, neurodegenerative outcomes are likely subject to ascertainment bias where those with COVID-19 have more contact with health services and thus are more likely to have neurodegenerative outcomes recorded, particularly in the early periods of follow-up. This may underpin the particularly high HRs observed initially (particularly on ‘day 0’), especially in those hospitalised when service contact is likely highest. We were only able to define cognitive impairment using codes from primary care, so the date of diagnosis for those initially identified in secondary care will be later and those identified in secondary care only will be missing. However, cognitive impairment is primarily managed in primary care with referrals to secondary care as needed, so we do not anticipate this will have had a large impact on outcome ascertainment. We could only assess COVID-19 severity according to hospitalization and did not consider the potential role of repeated infections. Our exposure was defined as diagnosed COVID-19, rather than SARS-CoV-2 infection, due to the lack of tests available at the start of the pandemic. This means that people who were asymptomatic or otherwise did not test or seek healthcare due to their infection were not considered to be exposed in our analyses. In addition, the effect of different COVID-19 variants on subsequent outcomes could not be directly accounted for as individual-level information on variant was not available, although the aligning of the cohorts to periods when specific variants were dominant in the population may have helped mitigate this effect (e.g. the pre-vaccination cohort is predominantly capturing the alpha variant). It is also difficult to capture interactions with healthcare using routinely collected data. We included a covariate that reflects GP practice visits in 2019 (‘consultation rate’) but our requirements for 6 months registration at a practice prior to index date may lead to an undercounting of GP practice visits in 2019 if patients have moved practice prior before index. Finally, we cannot exclude the possibility of unmeasured confounding, although we controlled for a wide range of demographic characteristics and prior morbidities.

Our findings highlight the consequences of COVID-19 on cognitive impairment, dementia and other neurodegenerative and neurological disorders in a dataset of more than 18 million individuals. In particular, we show that incidence of cognitive impairment may be elevated for years after COVID-19, especially amongst people who were unvaccinated when they were diagnosed with COVID-19, plus incidence of both cognitive impairment and dementia was generally more elevated after hospitalised COVID-19, compared with non-hospitalised COVID-19. These findings suggest a potential longer-term impact of COVID-19 on cognitive health, particularly among individuals unvaccinated at the time of COVID-19 diagnosis and/or who were hospitalised with COVID-19. Among such individuals, there is a need for awareness of possible cognitive sequelae during routine care and consideration of further evaluation.

## Data Availability

All non-disclosive data produced in the present work are contained in the manuscript. Access to the underlying identifiable and potentially re-identifiable pseudonymised electronic health record data is tightly governed by various legislative and regulatory frameworks, and restricted by best practice. The data in OpenSAFELY is drawn from General Practice data across England where TPP is the data processor. TPP developers initiate an automated process to create pseudonymised records in the core OpenSAFELY database, which are copies of key structured data tables in the identifiable records. These pseudonymised records are linked onto key external data resources that have also been pseudonymised via SHA-512 one-way hashing of NHS numbers using a shared salt. Bennett Institute for Applied Data Science developers and PIs holding contracts with NHS England have access to the OpenSAFELY pseudonymised data tables as needed to develop the OpenSAFELY tools. These tools in turn enable researchers with OpenSAFELY data access agreements to write and execute code for data management and data analysis without direct access to the underlying raw pseudonymised patient data, and to review the outputs of this code. All code for the full data management pipeline - from raw data to completed results for this analysis - and for the OpenSAFELY platform as a whole is available for review at https://github.com/OpenSAFELY.

## AUTHORS CONTRIBUTIONS STATEMENT

Author contributions are reported below in line with the Contributor Roles Taxonomy (CRediT). Conceptualization: Jonathan Sterne, Rachel Denholm, Elizabeth Coulthard, Venexia Walker Data curation: Harry T. Mason, Mengxuan Zou, Emily Eyles, Jose Ignacio Cuitun Coronado, Paul Madley-Dowd, Marwa Al Arab, Yinghui Wei, Genevieve Cezard, Tom Palmer, Seb Bacon, Simon Davy, Louis Fisher, Ben Goldacre, Rachel Denholm, Venexia Walker Formal analysis: Harry T. Mason, Mengxuan Zou, Emily Eyles, Jose Ignacio Cuitun Coronado, Marwa Al Arab, Yinghui Wei, Genevieve Cezard, Tom Palmer, Rachel Denholm, Venexia Walker Funding acquisition: Ben Goldacre, Nishi Chaturvedi, Jonathan Sterne Investigation: Harry T. Mason, Mengxuan Zou, Emily Eyles, Jose Ignacio Cuitun Coronado, Marwa Al Arab, Yinghui Wei, Genevieve Cezard, Tom Palmer, Rachel Denholm, Venexia Walker Methodology: Tom Palmer, Jonathan Sterne, Rachel Denholm, Venexia Walker Project administration: Amelia Green, Alex Walker, Ben Goldacre, Jonathan Sterne, Rachel Denholm, Venexia Walker Resources: Seb Bacon, Ben Goldacre

Software: Harry T. Mason, Mengxuan Zou, Emily Eyles, Jose Ignacio Cuitun Coronado, Marwa Al Arab, Yinghui Wei, Genevieve Cezard, Tom Palmer, Amelia Green, Alex Walker, Seb Bacon, Simon Davy, Louis Fisher, Ben Goldacre, Rachel Denholm, Venexia Walker Supervision: Amelia Green, Alex Walker, Louis Fisher, Amir Mehrkar, Ben Goldacre, Nishi Chaturvedi, Jonathan Sterne, Rachel Denholm, Elizabeth Coulthard, Venexia Walker Validation: Harry T. Mason, Mengxuan Zou, Emily Eyles, Jose Ignacio Cuitun Coronado, Marwa Al Arab, Yinghui Wei, Genevieve Cezard, Rachel Denholm, Venexia Walker Visualization: Harry T. Mason, Mengxuan Zou, Emily Eyles, Venexia Walker Writing - Original Draft: Harry T. Mason, Venexia Walker Writing - Review & Editing: All authors

## DATA SHARING STATEMENT

Access to the underlying identifiable and potentially re-identifiable pseudonymised electronic health record data is tightly governed by various legislative and regulatory frameworks, and restricted by best practice. The data in OpenSAFELY is drawn from General Practice data across England where TPP is the data processor. TPP developers initiate an automated process to create pseudonymised records in the core OpenSAFELY database, which are copies of key structured data tables in the identifiable records. These pseudonymised records are linked onto key external data resources that have also been pseudonymised via SHA-512 one-way hashing of NHS numbers using a shared salt. Bennett Institute for Applied Data Science developers and PIs holding contracts with NHS England have access to the OpenSAFELY pseudonymised data tables as needed to develop the OpenSAFELY tools. These tools in turn enable researchers with OpenSAFELY data access agreements to write and execute code for data management and data analysis without direct access to the underlying raw pseudonymised patient data, and to review the outputs of this code. All code for the full data management pipeline—from raw data to completed results for this analysis—and for the OpenSAFELY platform as a whole is available for review at https://github.com/OpenSAFELY.

## ACKNOWLEDGEMENTS

We are very grateful for all the support received from the TPP Technical Operations team throughout this work, and for generous assistance from the information governance and database teams at NHS England and the NHS England Transformation Directorate. We acknowledge members of the OpenSAFELY Collaborative, who have contributed to the development and maintenance of the OpenSAFELY platform. We thank the CONVALESCENCE Study Long COVID PPIE group for their input and for sharing their experiences and expertise throughout the duration of the project. Emily Eyles’ time is supported by NIHR ARC South West. The views expressed in this publication are those of the authors and not necessarily those of the NIHR or the Department of Health and Social Care.

## INFORMATION GOVERNANCE AND ETHICAL APPROVAL

NHS England is the data controller for OpenSAFELY-TPP; TPP is the data processor; all study authors using OpenSAFELY have the approval of NHS England. This implementation of OpenSAFELY is hosted within the TPP environment which is accredited to the ISO 27001 information security standard and is NHS IG Toolkit compliant (https://digital.nhs.uk/data-and-information/looking-after-information/data-security-and-information-governance/data-security-and-protection-toolkit).

Patient data are pseudonymised for analysis and linkage using industry standard cryptographic hashing techniques; all pseudonymised datasets transmitted for linkage onto OpenSAFELY are encrypted; access to the platform is via a virtual private network (VPN) connection, restricted to a small group of researchers; the researchers hold contracts with NHS England and only access the platform to initiate database queries and statistical models; all database activity is logged; only aggregate statistical outputs leave the platform environment following best practice for anonymisation of results such as statistical disclosure control for low cell counts (https://digital.nhs.uk/data-and-information/information-standards/information-standards-and-data-collections-including-extractions/publications-and-notifications/standards-and-collections/isb1523-anonymisation-standard-for-publishing-health-and-social-care-data).

The OpenSAFELY research platform adheres to the obligations of the UK General Data Protection Regulation (GDPR) and the Data Protection Act 2018. In March 2020, the Secretary of State for Health and Social Care used powers under the UK Health Service (Control of Patient Information) Regulations 2002 (COPI) to require organisations to process confidential patient information for the purposes of protecting public health, providing healthcare services to the public and monitoring and managing the COVID-19 outbreak and incidents of exposure; this sets aside the requirement for patient consent (https://web.archive.org/web/20200421171727/https://www.gov.uk/government/publications/covid-19-notification-to-gps-and-nhs-england-to-share-information). This was extended in November 2022 for the NHS England OpenSAFELY COVID-19 research platform (https://www.gov.uk/government/publications/coronavirus-covid-19-notification-to-organisations-to-share-information/coronavirus-covid-19-notice-under-regulation-34-of-the-health-service-control-of-patient-information-regulations-2002). In some cases of data sharing, the common law duty of confidence is met using, for example, patient consent or support from the Health Research Authority Confidentiality Advisory Group (https://www.hra.nhs.uk/about-us/committees-and-services/confidentiality-advisory-group/).

Taken together, these provide the legal bases to link patient datasets on the OpenSAFELY platform. GP practices, from which the primary care data are obtained, are required to share relevant health information to support the public health response to the pandemic, and have been informed of the OpenSAFELY analytics platform.

