## Supplement for "Association of COVID-19 with neurodegenerative and neurological disorders in vaccinated and unvaccinated people"

Supplementary Figure 10 Maximally adjusted hazard ratios and 95% CIs for Parkinson's disease and associated risk conditions following diagnosis of COVID-19 by cohort, overall, and by COVID-19 severity... 35

**Supplementary Text 1: Absolute excess risk**

The absolute excess risk analysis was performed for each outcome in each cohort using the hazard ratio from the main analysis. To compare the outcomes across the cohorts, each of which have different lengths of follow-up, we calculated the absolute excess risk at 52 weeks. We accounted for age and sex in the analysis by using event counts and person days from eight subgroups defined by age and sex. They were females, aged 18-49; females, aged 50-64; females, aged 65-84; females, aged 85-110; males, aged 18-49; males, aged 50-64; males, aged 65-84; and males, aged 85-110). For each group, we calculated the average daily incidence of the outcome in the unexposed and the cumulative risk over time. We then used the relevant hazard ratio for that day (e.g., days 1 to 27 will have the coefficient for the term 'days1_28', while days 28 to 182 will have the coefficient for the term 'days28_183') to predict the expected cumulative survival in the exposed. Finally, we calculated the daily excess risk as the difference in cumulative survival for the unexposed and the expected cumulative survival in the exposed. The overall absolute excess risk was estimated using a weighted sum of the excess risks in each group, weighted by the proportions of individuals in age and sex strata in the pre-vaccination cohort. Day 0 was excluded from the analysis.

**Supplementary Text 2: Deviations from protocol**

The following alterations were made from the pre-specified protocol (https://github.com/opensafely/post-covid-neurodegenerative):

- The following subgroups were restricted to only include dementia and cognitive impairment as outcomes due to low event counts: COVID-19 severity, age group, sex, ethnicity, and high vascular risk.
- If there were less than 12 events in any of the first three time periods (i.e. 0-4 weeks, 5-26 weeks, 27-52 weeks), we used a single time period (i.e., 1 year) instead. We also collapsed the 2-3 and 3-4 year periods into 2-4 years. The full set of outputs, including the models without the collapsed time periods, can be found at https://jobs.opensafely.org/investigating-events-following-covid-19/post-covid-neurodegenerative-v3/outputs. Models with low counts should be interpreted with extreme caution.
- An error in the original protocol listed Wales and Scotland as regions for analysis, as the protocol states elsewhere that only English GP data will be included. Additionally, the "Midlands" region was separated into "East Midlands" and "West Midlands", and the "North East and Yorkshire" region was separated into "North East" and "Yorkshire and the Humber".
- The initial protocol used CTV3 labels for ethnicity, which were updated to fit with the SNOMED-CT codelists combined with SUS secondary care data that were used for this work ("South Asian" became "Asian or Asian British", "Black" became "Black or Black British", "Other" became "Chinese or Other Ethnic Group").
- The age groups 18-39 and 40-64 were updated to be 18-49 and 50-64 to better reflect clinically important ages for neurodegenerative outcomes.
- The outcomes Parkinson's disease and dementia (including its subtypes) were excluded for the age group 18-49 due to low event counts.
- The end date was extended from April 30th 2024 to May 31st 2025, to allow for the increased data available at runtime. Cox regression time periods were also updated accordingly.
- People with outcomes on 'day 0' were removed from the analysis.

**Supplementary Table 1: Summary of covariate definitions**

| Covariate | Type | Definition |
|---|---|---|
| **Age** | Continuous | Modelled as age in years using a restricted cubic spline with 3 knots at the 10th, 50th and 90th percentiles |
| **Sex** | Categorical | Male, Female |
| **Ethnicity** | Categorical | 1: White<br>2: Mixed<br>3: Asian or Asian British<br>4: Black or Black British<br>5: Chinese or Other Ethnic Groups |
| **Deprivation** | Categorical | Index of Multiple Deprivation 2019 deciles |
| **Region** | Categorical | East of England<br>East Midlands<br>London<br>North East<br>North West<br>South East<br>South West<br>West Midlands<br>Yorkshire and the Humber |
| **Smoking status** | Categorical | E: Ever smoker<br>M: Missing<br>N: Never smoker<br>S: Current smoker |
| **Care home status** | Binary | 1 if care home resident; 0 otherwise |
| **Consultation rate** | Continuous | Number of GP consultations in 2019 (i.e., annual consultations prior to the pandemic) |
| **Health care worker** | Binary | 1 if healthcare worker; 0 otherwise |
| **Liver disease** | Binary | 1 if diagnosis present; 0 otherwise. |
| **Chronic kidney disease** | Binary | 1 if diagnosis present; 0 otherwise. |
| **Cancer** | Categorical | 1 if diagnosis present; 0 otherwise. |
| **Hypertension** | Binary | 1 if diagnosis present; 0 otherwise. |
| **Diabetes** | Binary | 1 if diagnosis present; 0 otherwise. |
| **Obesity** | Binary | 1 if BMI>=30 or coded diagnosis for obesity; 0 otherwise. |
| **Chronic obstructive pulmonary disease (COPD)** | Binary | 1 if diagnosis present; 0 otherwise. |
| **Acute myocardial infarction** | Binary | 1 if diagnosis present; 0 otherwise. |
| **Ischaemic stroke** | Binary | 1 if diagnosis present; 0 otherwise. |
| **Depression** | Binary | 1 if diagnosis present; 0 otherwise |
| **History of cognitive impairment*** | Binary | 1 if diagnosis present; 0 otherwise |
| **History of dementia*** | Binary | 1 if diagnosis present; 0 otherwise |
| **History of migraine*** | Binary | 1 if diagnosis present; 0 otherwise |
| **History of motor neurone disease*** | Binary | 1 if diagnosis present; 0 otherwise |
| **History of multiple sclerosis*** | Binary | 1 if diagnosis present; 0 otherwise |
| **History of Parkinson's disease*** | Binary | 1 if diagnosis present; 0 otherwise |
| **History of Parkinson's risk conditions*†** | Binary | 1 if diagnosis present; 0 otherwise |

*The covariate "history of X" will not be included in the model when investigating the outcomes involving "X"

†Parkinson's risk conditions: REM sleep disorder and/or restless leg syndrome

**Supplementary Table 2: Summary of cohort characteristics**

| | Cohort 1:<br>Pre-vaccination | Cohort 2:<br>Vaccinated | Cohort 3:<br>Unvaccinated |
|---|---|---|---|
| **Start date** | January 1st 2020, which is the approximate start date of the pandemic in the UK. | June 1st 2021, the date that the delta variant was thought to be ubiquitous in England. | June 1st 2021, the date that the delta variant was thought to be ubiquitous in England. |
| **End date - exposure** | June 18th 2021, the date when the Joint Committee for Vaccination and Immunisation (JCVI) phase 2, group 12 (all adults aged 18 years and older) become eligible for a COVID-19 vaccination. | December 14th 2021, the day that the UK Health Security Agency stated that over half of English cases they sampled have S Gene Target Failure, meaning they were likely Omicron*. | December 14th 2021, the day that the UK Health Security Agency stated that over half of English cases they sampled have S Gene Target Failure, meaning they were likely Omicron*. |
| **End date - outcome** | May 31st 2025 | May 31st 2025 | May 31st 2025 |
| **Exclusion criteria** | Patients will be excluded if they meet any of the following criteria:<br>• COVID-19 diagnosis recorded prior to their index date | Patients will be excluded if they meet any of the following criteria:<br>• COVID-19 diagnosis recorded prior to their index date<br>• They do not have a record of two vaccination doses prior to the study end date<br>• They received a vaccination prior to December 8th 2020 (i.e., the start of the vaccination program)<br>• They received a second dose vaccination before their first dose vaccination<br>• They received a second dose vaccination less than three weeks after their first dose<br>• They received mixed vaccine products before May 7th 2021 | Patients will be excluded if they meet any of the following criteria:<br>• COVID-19 diagnosis recorded prior to their index date<br>• They have a record of one or more vaccination doses prior to their index date<br>• They could not be assigned to a vaccination group as defined by the Joint Committee on Vaccination and Immunisation (JCVI) |
| **Follow-up start (i.e., an individual's index date)** | Study start date. | Follow-up will start at the latest of the following dates:<br>• Two weeks after their second vaccination<br>• Study start date | Follow-up will start at the latest of the following dates:<br>• 12 weeks after they became eligible for vaccination<br>• Study start date |
| **Follow-up end** | Follow-up will end at the earliest of the following dates:<br>• Death<br>• Outcome event<br>• Study end date outcome<br>• Deregistration date | Follow-up will end at the earliest of the following dates:<br>• Death<br>• Outcome event<br>• Study end date outcome<br>• Deregistration date | Follow-up will end at the earliest of the following dates:<br>• Death<br>• Outcome event<br>• Study end date outcome<br>• Deregistration date |

| | | | |
|---|---|---|---|
| **Cox regression time periods, full** | [1,28), [28,183), [183, 365), [365,730), [730,1095), [1095,1460), [1460,1979) | [1,28), [28,183), [183, 365), [365,730), [730,1095), [1095,1460) | [1,28), [28,183), [183, 365), [365,730), [730,1095), [1095,1460) |
| **Cox regression time periods, collapsed** | [1, 365), [365,730), [730,1460), [1460,1979) | [1, 365), [365,730), [730, 1460) | [1, 365), [365,730), [730, 1460) |

*** UK Health Security Agency. (2021). Omicron daily overview: December 17th 2021. Retrieved April 26th 2024, from [https://assets.publishing.service.gov.uk/government/uploads/system/uploads/attachment_data/file/1042100/20211217_OS_Daily_Omicron_Overview.pdf](https://assets.publishing.service.gov.uk/government/uploads/system/uploads/attachment_data/file/1042100/20211217_OS_Daily_Omicron_Overview.pdf)**

**Supplementary Table 3: Patient medical history by cohort.**

| Characteristics | Pre-vaccination cohort | | Vaccinated cohort | | Unvaccinated cohort | |
|---|---|---|---|---|---|---|
| | N (%) | COVID-19 diagnoses | N (%) | COVID-19 diagnoses | N (%) | COVID-19 diagnoses |
| Total, No. | 18,334,845 | 954,945 | 13,805,541 | 850,581 | 3,185,973 | 144,273 |
| Acute myocardial infarction | 460,179 (2.5%) | 21,753 | 417,759 (3.0%) | 16,233 | 21,027 (0.7%) | 1,071 |
| Cancer | 5,328,153 (29.1%) | 315,489 | 4,650,999 (33.7%) | 353,481 | 586,449 (18.4%) | 47,577 |
| Cognitive impairment | 587,289 (3.2%) | 35,781 | 501,039 (3.6%) | 22,125 | 42,723 (1.3%) | 2,523 |
| Chronic kidney disease | 1,156,497 (6.3%) | 53,385 | 1,052,901 (7.6%) | 37,347 | 46,155 (1.4%) | 2,523 |
| Chronic obstructive pulmonary disorder (COPD) | 684,561 (3.7%) | 31,059 | 606,969 (4.4%) | 24,057 | 34,029 (1.1%) | 1,647 |
| Dementia | 217,641 (1.2%) | 20,043 | 164,937 (1.2%) | 5,511 | 5,523 (0.2%) | 285 |
| Depression | 5,224,983 (28.5%) | 289,011 | 4,241,109 (30.7%) | 280,695 | 705,855 (22.2%) | 46,707 |
| Diabetes | 1,677,879 (9.2%) | 92,439 | 1,661,313 (12.0%) | 85,137 | 126,699 (4.0%) | 7,611 |
| Hypertension | 6,261,579 (34.2%) | 302,607 | 5,472,885 (39.6%) | 283,035 | 47,6673 (15%) | 30,243 |
| Liver disease | 133,119 (0.7%) | 6,669 | 115,857 (0.8%) | 5,625 | 16,089 (0.5%) | 705 |
| Migraine | 1,645,653 (9.0%) | 102,393 | 1,376,847 (10%) | 102,933 | 190,683 (6.0%) | 15,183 |
| Motor neurone disease | 3,573 (0.0%) | 165 | 3,171 (0.0%) | 129 | 225 (0.0%) | 9 |
| Multiple sclerosis | 57,543 (0.3%) | 2,739 | 51,783 (0.4%) | 3,063 | 4,131 (0.1%) | 261 |
| Obesity | 1,493,433 (8.1%) | 91,797 | 1,462,413 (10.6%) | 98,943 | 145,899 (4.6%) | 11,841 |
| Parkinson's disease | 56,637 (0.3%) | 3363 | 48,459 (0.4%) | 1,527 | 1,515 (0.0%) | 63 |
| Restless leg syndrome and/or REM sleep disorder | 234,513 (1.3%) | 13,113 | 226,893 (1.6%) | 13,251 | 15,063 (0.5%) | 1,137 |
| Ischaemic stroke | 297,129 (1.6%) | 14,751 | 269,145 (1.9%) | 9,501 | 13,509 (0.4%) | 687 |

**Supplementary Table 4: Sex- and age-adjusted hazard ratios and 95% confidence intervals for cognitive impairment and dementia following diagnosis of COVID-19 by cohort following: any diagnosis of COVID-19, diagnosis of hospitalised COVID-19, and diagnosis of non-hospitalised COVID-19**

| Outcome | Time since COVID-19 | Pre-vaccination cohort | Vaccinated cohort | Unvaccinated cohort |
|---|---|---|---|---|
| **All COVID-19** | | | | |
| | 0-4 weeks | 4.89 (4.55-5.25) | 2.37 (2.17-2.59) | 5.65 (4.45-7.16) |
| | 5-26 weeks | 2.37 (2.27-2.48) | 1.46 (1.39-1.53) | 2.44 (2.09-2.85) |
| | 27-52 weeks | 1.62 (1.55-1.70) | 1.08 (1.02-1.13) | 1.91 (1.63-2.23) |
| **Cognitive impairment** | 1-2 years | 1.37 (1.33-1.42) | 1.00 (0.97-1.04) | 1.76 (1.58-1.97) |
| | 2-3 years | 1.23 (1.18-1.27) | 1.11 (1.07-1.14) | 2.01 (1.82-2.21) |
| | 3-4 years | 1.25 (1.21-1.30) | 1.21 (1.17-1.26) | 1.77 (1.57-2.00) |
| | 4-5.5 years | 1.43 (1.37-1.49) | - | - |
| | 0-4 weeks | 6.47 (5.93-7.07) | 2.22 (1.92-2.57) | 12.97 (9.23-18.21) |
| | 5-26 weeks | 2.53 (2.37-2.70) | 1.21 (1.11-1.32) | 2.03 (1.39-2.97) |
| | 27-52 weeks | 1.79 (1.66-1.92) | 1.03 (0.94-1.12) | 1.99 (1.37-2.91) |
| **Dementia** | 1-2 years | 1.43 (1.35-1.51) | 0.94 (0.88-1.00) | 1.58 (1.16-2.16) |
| | 2-3 years | 1.30 (1.23-1.39) | 0.93 (0.87-0.99) | 1.35 (0.95-1.92) |
| | 3-4 years | 1.29 (1.21-1.37) | 0.90 (0.83-0.98) | 1.79 (1.23-2.61) |
| | 4-5.5 years | 1.21 (1.11-1.32) | - | - |
| **Hospitalised COVID-19** | | | | |
| | 0-4 weeks | 21.41 (19.88-23.05) | 15.14 (13.64-16.81) | 13.94 (9.13-21.28) |
| | 5-26 weeks | 4.72 (4.27-5.22) | 3.71 (3.19-4.32) | 5.84 (4.30-7.93) |
| | 27-52 weeks | 2.82 (2.51-3.17) | 2.86 (2.43-3.36) | 3.03 (2.07-4.43) |
| **Cognitive impairment** | 1-2 years | 2.22 (2.03-2.43) | 1.86 (1.61-2.15) | 2.57 (1.93-3.43) |
| | 2-3 years | 1.86 (1.68-2.05) | 1.96 (1.69-2.27) | 2.45 (1.87-3.22) |
| | 3-4 years | 1.96 (1.78-2.16) | 1.74 (1.44-2.12) | 2.23 (1.61-3.10) |
| | 4-5.5 years | 1.69 (1.49-1.92) | - | - |
| | 0-4 weeks | 19.69 (17.88-21.67) | 20.29 (18.24-22.58) | |
| | 5-26 weeks | 3.81 (3.36-4.31) | 2.88 (2.38-3.49) | 1.70 (1.21-2.39) |
| | 27-52 weeks | 2.59 (2.24-2.98) | 2.52 (2.06-3.08) | |
| **Dementia** | 1-2 years | 2.18 (1.94-2.44) | 1.69 (1.41-2.03) | 1.42 (0.96-2.11) |
| | 2-3 years | 1.77 (1.55-2.02) | 1.68 (1.38-2.04) | 1.19 (0.84-1.68) |
| | 3-4 years | 1.74 (1.52-2.00) | 1.53 (1.18-1.98) | |
| | 4-5.5 years | 1.66 (1.39-1.98) | - | - |
| **Non-hospitalised COVID-19** | | | | |
| | 0-4 weeks | 3.91 (3.59-4.26) | 2.13 (1.93-2.34) | 4.42 (3.33-5.88) |
| | 5-26 weeks | 2.12 (2.02-2.23) | 1.37 (1.30-1.44) | 2.02 (1.69-2.41) |
| | 27-52 weeks | 1.51 (1.43-1.58) | 1.01 (0.96-1.07) | 1.76 (1.49-2.09) |
| **Cognitive impairment** | 1-2 years | 1.29 (1.25-1.34) | 0.98 (0.94-1.01) | 1.65 (1.47-1.87) |
| | 2-3 years | 1.17 (1.13-1.21) | 1.08 (1.05-1.12) | 1.94 (1.75-2.15) |
| | 3-4 years | 1.19 (1.15-1.24) | 1.20 (1.15-1.24) | 1.70 (1.50-1.93) |
| | 4-5.5 years | 1.41 (1.35-1.47) | - | - |
| | 0-4 weeks | 3.81 (3.34-4.33) | 1.24 (1.01-1.53) | |
| | 5-26 weeks | 2.25 (2.09-2.43) | 1.06 (0.96-1.16) | 5.53 (4.21-7.26) |
| | 27-52 weeks | 1.62 (1.49-1.76) | 0.90 (0.82-0.99) | |
| **Dementia** | 1-2 years | 1.28 (1.20-1.37) | 0.89 (0.83-0.95) | 1.95 (1.19-3.20) |
| | 2-3 years | 1.22 (1.13-1.30) | 0.88 (0.82-0.94) | 2.28 (1.57-3.31) |
| | 3-4 years | 1.21 (1.12-1.30) | 0.86 (0.79-0.94) | |
| | 4-5.5 years | 1.12 (1.02-1.24) | - | - |

- Time period falls outside of cohort follow-up

**Supplementary Table 5: Maximally adjusted hazard ratios and 95% CIs for cognitive impairment and dementia following diagnosis of COVID-19 in the vaccinated cohort in people with a prior history of COVID-19.**

| Outcome | Time since COVID-19 | Vaccinated cohort |
|---|---|---|
| Cognitive impairment | 0-4 weeks | 2.19 (1.39-3.44) |
| | 5-26 weeks | 0.92 (0.68-1.25) |
| | 27-52 weeks | 1.32 (1.04-1.68) |
| | 1-2 years | 0.96 (0.78-1.18) |
| | 2-3 years | 0.79 (0.63-0.99) |
| | 3-4 years | 0.85 (0.66-1.08) |
| Dementia | 0-1 years | 1.06 (0.80-1.41) |
| | 1-2 years | 0.83 (0.58-1.19) |
| | 2-4 years | 1.10 (0.85-1.43) |

**Supplementary Table 6: Maximally adjusted hazard ratios and 95% CIs for cognitive impairment following diagnosis of COVID-19 and by age group.**

| Outcome | Time since COVID-19 | Pre-vaccination cohort | Vaccinated cohort | Unvaccinated cohort |
|---|---|---|---|---|
| | 0-4 weeks | 4.16 (3.58-4.84) | 3.21 (2.73-3.77) | 3.83 (2.73-5.39) |
| | 5-26 weeks | 2.36 (2.17-2.57) | 1.73 (1.58-1.90) | 1.85 (1.50-2.28) |
| | 27-52 weeks | 1.62 (1.49-1.77) | 1.06 (0.96-1.17) | 1.56 (1.28-1.91) |
| **18-49 years** | 1-2 years | 1.24 (1.16-1.32) | 1.02 (0.96-1.10) | 1.35 (1.17-1.57) |
| | 2-3 years | 1.11 (1.04-1.19) | 1.04 (0.98-1.10) | 1.48 (1.32-1.67) |
| | 3-4 years | 1.03 (0.97-1.09) | 1.09 (1.02-1.15) | 1.30 (1.13-1.51) |
| | 4-5.5 years | 1.11 (1.04-1.19) | - | - |
| | 0-4 weeks | 4.60 (3.96-5.35) | 2.71 (2.30-3.20) | 6.02 (3.84-9.45) |
| | 5-26 weeks | 1.99 (1.82-2.19) | 1.51 (1.37-1.66) | 1.93 (1.38-2.70) |
| | 27-52 weeks | 1.59 (1.46-1.74) | 1.02 (0.93-1.13) | 1.59 (1.15-2.21) |
| **50-64 years** | 1-2 years | 1.27 (1.19-1.36) | 0.97 (0.91-1.04) | 1.35 (1.06-1.71) |
| | 2-3 years | 1.09 (1.02-1.16) | 1.00 (0.94-1.07) | 1.42 (1.13-1.77) |
| | 3-4 years | 1.12 (1.05-1.19) | 1.01 (0.93-1.09) | 1.27 (0.96-1.67) |
| | 4-5.5 years | 1.15 (1.06-1.25) | - | - |
| | 0-4 weeks | 4.92 (4.39-5.52) | 1.75 (1.48-2.06) | 5.96 (3.50-10.16) |
| | 5-26 weeks | 2.01 (1.86-2.17) | 1.23 (1.13-1.34) | 2.32 (1.56-3.46) |
| | 27-52 weeks | 1.27 (1.17-1.39) | 1.06 (0.97-1.15) | 1.51 (0.98-2.31) |
| **65-84 years** | 1-2 years | 1.20 (1.13-1.28) | 0.98 (0.92-1.04) | 1.45 (1.05-2.02) |
| | 2-3 years | 1.10 (1.03-1.17) | 1.01 (0.95-1.07) | 1.62 (1.18-2.21) |
| | 3-4 years | 1.09 (1.02-1.16) | 0.98 (0.92-1.05) | 1.37 (0.91-2.08) |
| | 4-5.5 years | 1.15 (1.06-1.25) | - | - |
| | 0-4 weeks | 2.61 (2.14-3.18) | 2.01 (1.56-2.59) | † |
| | 5-26 weeks | 1.84 (1.64-2.06) | 1.40 (1.21-1.62) | † |
| | 27-52 weeks | 1.21 (1.05-1.38) | 1.18 (1.01-1.38) | † |
| **85-110 years** | 1-2 years | 1.14 (1.02-1.27) | 1.01 (0.89-1.14) | † |
| | 2-3 years | 1.24 (1.09-1.41) | 0.91 (0.79-1.06) | † |
| | 3-4 years | 0.94 (0.79-1.13) | 0.93 (0.76-1.15) | † |
| | 4-5.5 years | 1.22 (0.98-1.52) | - | - |

† Insufficient events for estimation - Time period falls outside of cohort follow-up

**Supplementary Table 7: Maximally adjusted hazard ratios and 95% CIs for dementia following diagnosis of COVID-19 and by age group.**

| Outcome | Time since COVID-19 | Pre-vaccination cohort | Vaccinated cohort | Unvaccinated cohort |
|---|---|---|---|---|
| 50-64 years | 0-4 weeks | 5.24 (3.69-7.46) | 0.75 (0.58-0.96) | † |
| | 5-26 weeks | 1.17 (0.86-1.60) | | † |
| | 27-52 weeks | 0.93 (0.69-1.26) | | † |
| | 1-2 years | 1.18 (0.98-1.42) | 0.83 (0.66-1.03) | † |
| | 2-3 years | 1.01 (0.84-1.22) | 0.87 (0.75-1.02) | † |
| | 3-4 years | 1.09 (0.92-1.29) | | † |
| | 4-5.5 years | 1.14 (0.91-1.41) | - | - |
| 65-84 years | 0-4 weeks | 5.67 (5.02-6.39) | 1.73 (1.39-2.14) | 10.78 (7.05-16.48) |
| | 5-26 weeks | 2.23 (2.04-2.43) | 1.02 (0.91-1.15) | 1.54 (0.92-2.58) |
| | 27-52 weeks | 1.58 (1.43-1.73) | 0.99 (0.88-1.10) | 1.62 (1.00-2.64) |
| | 1-2 years | 1.23 (1.14-1.32) | 0.91 (0.84-0.98) | 1.00 (0.64-1.56) |
| | 2-3 years | 1.19 (1.11-1.28) | 0.90 (0.84-0.97) | 1.17 (0.76-1.79) |
| | 3-4 years | 1.12 (1.04-1.21) | 0.84 (0.77-0.93) | 0.99 (0.55-1.76) |
| | 4-5.5 years | 1.02 (0.92-1.13) | - | - |
| 85-110 years | 0-4 weeks | 4.96 (4.32-5.69) | 3.12 (2.53-3.85) | 2.55 (1.71-3.80) |
| | 5-26 weeks | 2.06 (1.85-2.29) | 1.64 (1.43-1.88) | |
| | 27-52 weeks | 1.51 (1.33-1.70) | 1.17 (0.99-1.37) | |
| | 1-2 years | 1.23 (1.10-1.37) | 1.03 (0.90-1.17) | 1.49 (0.80-2.77) |
| | 2-3 years | 1.05 (0.91-1.22) | 0.99 (0.85-1.16) | 1.76 (1.00-3.10) |
| | 3-4 years | 1.20 (1.01-1.42) | 0.93 (0.75-1.14) | |
| | 4-5.5 years | 1.08 (0.85-1.37) | - | - |

† Insufficient events for estimation - Time period falls outside of cohort follow-up

**Supplementary Table 8: Maximally adjusted hazard ratios and 95% CIs for cognitive impairment and dementia following diagnosis of COVID-19 by cohort and by sex.**

| Outcome | Time since COVID-19 | Pre-vaccination cohort | Vaccinated cohort | Unvaccinated cohort |
|---|---|---|---|---|
| **Cognitive impairment** | | | | |
| **Women** | 0-4 weeks | 3.53 (3.19-3.91) | 2.32 (2.07-2.61) | 3.69 (2.64-5.15) |
| | 5-26 weeks | 2.04 (1.93-2.16) | 1.39 (1.31-1.49) | 1.96 (1.61-2.39) |
| | 27-52 weeks | 1.41 (1.32-1.49) | 1.03 (0.96-1.10) | 1.60 (1.32-1.93) |
| | 1-2 years | 1.17 (1.12-1.23) | 0.93 (0.89-0.97) | 1.35 (1.17-1.56) |
| | 2-3 years | 1.09 (1.04-1.14) | 1.07 (1.03-1.12) | 1.45 (1.28-1.64) |
| | 3-4 years | 1.15 (1.10-1.20) | 1.23 (1.17-1.29) | 1.31 (1.13-1.52) |
| | 4-5.5 years | 1.35 (1.29-1.43) | - | - |
| **Men** | 0-4 weeks | 4.68 (4.22-5.19) | 2.11 (1.83-2.42) | 5.74 (4.09-8.05) |
| | 5-26 weeks | 1.94 (1.81-2.08) | 1.40 (1.30-1.50) | 1.85 (1.42-2.40) |
| | 27-52 weeks | 1.38 (1.28-1.49) | 1.04 (0.96-1.12) | 1.28 (0.97-1.70) |
| | 1-2 years | 1.23 (1.17-1.30) | 0.99 (0.94-1.05) | 1.41 (1.17-1.69) |
| | 2-3 years | 1.10 (1.04-1.16) | 1.02 (0.97-1.08) | 1.74 (1.49-2.04) |
| | 3-4 years | 1.11 (1.05-1.17) | 1.02 (0.96-1.09) | 1.44 (1.17-1.78) |
| | 4-5.5 years | 1.16 (1.08-1.24) | - | - |
| **Dementia** | | | | |
| **Women** | 0-4 weeks | 4.25 (3.74-4.82) | 1.96 (1.59-2.40) | 9.31 (6.06-14.29) |
| | 5-26 weeks | 2.03 (1.86-2.21) | 1.12 (0.99-1.26) | 1.47 (0.89-2.43) |
| | 27-52 weeks | 1.43 (1.30-1.58) | 0.94 (0.83-1.06) | 1.29 (0.77-2.16) |
| | 1-2 years | 1.16 (1.07-1.25) | 0.90 (0.82-0.98) | 1.17 (0.78-1.76) |
| | 2-3 years | 1.07 (0.98-1.16) | 0.85 (0.78-0.93) | 1.19 (0.77-1.83) |
| | 3-4 years | 1.16 (1.06-1.26) | 0.78 (0.69-0.88) | 1.35 (0.82-2.22) |
| | 4-5.5 years | 1.03 (0.91-1.15) | - | - |
| **Men** | 0-4 weeks | 6.11 (5.40-6.91) | 2.18 (1.77-2.69) | |
| | 5-26 weeks | 1.96 (1.77-2.18) | 1.17 (1.03-1.32) | 2.32 (1.62-3.32) |
| | 27-52 weeks | 1.45 (1.29-1.62) | 1.02 (0.89-1.15) | |
| | 1-2 years | 1.22 (1.12-1.33) | 0.91 (0.83-1.00) | 1.25 (0.76-2.07) |
| | 2-3 years | 1.19 (1.09-1.31) | 0.94 (0.86-1.03) | 1.11 (0.71-1.72) |
| | 3-4 years | 1.10 (0.99-1.21) | 0.96 (0.86-1.07) | |
| | 4-5.5 years | 1.10 (0.96-1.25) | - | - |

- Time period falls outside of cohort follow-up

**Supplementary Table 9: Maximally adjusted hazard ratios and 95% CIs for cognitive impairment following diagnosis of COVID-19 by cohort and by ethnicity**

| Outcome | Time since COVID-19 | Pre-vaccination cohort | Vaccinated cohort | Unvaccinated cohort |
|---|---|---|---|---|
| **White** | 0-4 weeks | 3.94 (3.64-4.25) | 2.19 (2.00-2.41) | 4.16 (3.19-5.43) |
| | 5-26 weeks | 2.00 (1.91-2.10) | 1.39 (1.33-1.47) | 1.80 (1.51-2.14) |
| | 27-52 weeks | 1.38 (1.32-1.45) | 1.03 (0.98-1.09) | 1.47 (1.24-1.75) |
| | 1-2 years | 1.18 (1.14-1.22) | 0.95 (0.92-0.99) | 1.33 (1.18-1.51) |
| | 2-3 years | 1.08 (1.04-1.12) | 1.05 (1.02-1.09) | 1.51 (1.36-1.69) |
| | 3-4 years | 1.12 (1.08-1.16) | 1.14 (1.10-1.19) | 1.30 (1.14-1.49) |
| | 4-5.5 years | 1.29 (1.23-1.35) | - | - |
| **Asian or Asian British** | 0-4 weeks | 4.48 (3.43-5.85) | 2.97 (1.98-4.45) | |
| | 5-26 weeks | 1.85 (1.56-2.20) | 1.41 (1.10-1.81) | 1.69 (1.11-2.57) |
| | 27-52 weeks | 1.38 (1.17-1.63) | 1.05 (0.81-1.36) | |
| | 1-2 years | 1.28 (1.14-1.44) | 1.10 (0.92-1.30) | 1.41 (0.93-2.16) |
| | 2-3 years | 1.11 (0.98-1.24) | 1.08 (0.92-1.28) | 1.66 (1.23-2.22) |
| | 3-4 years | 1.07 (0.95-1.19) | 1.11 (0.92-1.35) | |
| | 4-5.5 years | 1.15 (1.00-1.32) | - | - |
| **Black or Black British** | 0-1 years | 1.62 (1.27-2.07) | 1.26 (0.88-1.82) | 1.86 (1.18-2.94) |
| | 1-2 years | 1.22 (0.94-1.58) | 0.82 (0.54-1.25) | 1.23 (0.73-2.07) |
| | 2-4 years | 1.29 (1.09-1.53) | 1.15 (0.87-1.50) | 1.24 (0.86-1.78) |
| | 4-5.5 years | 1.33 (0.98-1.80) | - | - |
| **Mixed** | 0-1 years | 2.27 (1.71-3.00) | 0.70 (0.40-1.21) | † |
| | 1-2 years | 1.10 (0.77-1.57) | 0.78 (0.47-1.28) | † |
| | 2-4 years | 1.07 (0.83-1.36) | 1.22 (0.90-1.64) | † |
| | 4-5.5 years | 1.04 (0.68-1.60) | - | - |
| **Chinese or other** | 0-1 years | 2.44 (1.82-3.28) | 1.46 (1.00-2.16) | † |
| | 1-2 years | 1.34 (0.95-1.90) | 0.99 (0.63-1.57) | † |
| | 2-4 years | 1.23 (0.96-1.57) | 0.91 (0.63-1.32) | † |
| | 4-5.5 years | 1.85 (1.25-2.74) | - | - |

† Insufficient events for estimation - Time period falls outside of cohort follow-up

**Supplementary Table 10: Maximally adjusted hazard ratios and 95% CIs for dementia following diagnosis of COVID-19 by cohort and by ethnicity**

| Outcome | Time since COVID-19 | Pre-vaccination cohort | Vaccinated cohort | Unvaccinated cohort |
|---|---|---|---|---|
| **White** | 0-4 weeks | 4.87 (4.44-5.35) | 1.94 (1.66-2.27) | 6.99 (4.65-10.51) |
| | 5-26 weeks | 2.04 (1.90-2.18) | 1.14 (1.05-1.25) | 1.46 (0.97-2.20) |
| | 27-52 weeks | 1.45 (1.35-1.57) | 0.98 (0.90-1.08) | 1.34 (0.88-2.05) |
| | 1-2 years | 1.18 (1.11-1.26) | 0.90 (0.84-0.96) | 1.06 (0.74-1.52) |
| | 2-3 years | 1.11 (1.04-1.19) | 0.89 (0.83-0.95) | 0.93 (0.62-1.39) |
| | 3-4 years | 1.13 (1.05-1.21) | 0.85 (0.78-0.92) | 1.02 (0.63-1.65) |
| | 4-5.5 years | 1.04 (0.95-1.14) | - | - |
| **Asian or Asian British** | 0-4 weeks | 5.63 (3.86-8.23) | | † |
| | 5-26 weeks | 1.21 (0.85-1.71) | 1.02 (0.74-1.40) | † |
| | 27-52 weeks | 1.07 (0.77-1.49) | | † |
| | 1-2 years | 0.95 (0.75-1.20) | 1.02 (0.74-1.40) | † |
| | 2-3 years | 1.16 (0.93-1.45) | 0.92 (0.71-1.19) | † |
| | 3-4 years | 1.19 (0.96-1.47) | | † |
| | 4-5.5 years | 1.02 (0.75-1.38) | - | - |
| **Black or Black British** | 0-1 years | 2.61 (1.81-3.76) | † | † |
| | 1-2 years | 1.39 (0.86-2.24) | † | † |
| | 2-4 years | 0.79 (0.51-1.24) | † | † |
| | 4-5.5 years | 0.97 (0.47-1.98) | - | - |

† Insufficient events for estimation - Time period falls outside of cohort follow-up

**Supplementary Table 11 Maximally adjusted hazard ratios and 95% CIs for cognitive impairment and dementia following diagnosis of COVID-19 by cohort and by vascular risk (defined as history of hypertension and/or diabetes)**

| Outcome | Time since COVID-19 | Pre-vaccination cohort | Vaccinated cohort | Unvaccinated cohort |
|---|---|---|---|---|
| **Cognitive impairment** | | | | |
| **Prior history of high vascular risk** | 0-4 weeks | 3.56 (3.26-3.89) | 1.97 (1.76-2.20) | 3.03 (2.08-4.42) |
| | 5-26 weeks | 1.86 (1.76-1.96) | 1.28 (1.20-1.35) | 1.80 (1.45-2.23) |
| | 27-52 weeks | 1.31 (1.24-1.39) | 1.07 (1.01-1.13) | 1.33 (1.07-1.65) |
| | 1-2 years | 1.17 (1.12-1.22) | 0.93 (0.89-0.97) | 1.24 (1.06-1.46) |
| | 2-3 years | 1.07 (1.02-1.11) | 1.02 (0.97-1.06) | 1.39 (1.20-1.60) |
| | 3-4 years | 1.10 (1.05-1.15) | 1.05 (1.00-1.10) | 1.24 (1.04-1.49) |
| | 4-5.5 years | 1.23 (1.17-1.30) | - | - |
| **No prior history of high vascular risk** | 0-4 weeks | 5.29 (4.67-5.99) | 3.03 (2.62-3.51) | 5.96 (4.38-8.12) |
| | 5-26 weeks | 2.34 (2.17-2.53) | 1.73 (1.60-1.88) | 1.94 (1.54-2.44) |
| | 27-52 weeks | 1.58 (1.46-1.71) | 0.97 (0.88-1.06) | 1.60 (1.28-2.00) |
| | 1-2 years | 1.25 (1.18-1.33) | 1.03 (0.97-1.09) | 1.44 (1.22-1.69) |
| | 2-3 years | 1.11 (1.04-1.18) | 1.10 (1.04-1.16) | 1.65 (1.45-1.88) |
| | 3-4 years | 1.12 (1.06-1.18) | 1.25 (1.18-1.33) | 1.43 (1.22-1.69) |
| | 4-5.5 years | 1.28 (1.20-1.37) | - | - |
| **Dementia** | | | | |
| **Prior history of high vascular risk** | 0-4 weeks | 5.10 (4.65-5.60) | 2.25 (1.93-2.62) | 8.47 (5.78-12.41) |
| | 5-26 weeks | 2.00 (1.87-2.15) | 1.15 (1.05-1.26) | 1.57 (1.04-2.36) |
| | 27-52 weeks | 1.51 (1.39-1.63) | 1.00 (0.90-1.10) | 1.50 (0.99-2.29) |
| | 1-2 years | 1.21 (1.14-1.29) | 0.90 (0.84-0.97) | 1.26 (0.90-1.78) |
| | 2-3 years | 1.15 (1.07-1.23) | 0.91 (0.85-0.98) | 1.00 (0.67-1.51) |
| | 3-4 years | 1.14 (1.06-1.22) | 0.88 (0.81-0.97) | 1.44 (0.93-2.20) |
| | 4-5.5 years | 1.07 (0.97-1.18) | - | - |
| **No prior history of high vascular risk** | 0-4 weeks | 4.91 (3.74-6.44) | 1.06 (0.63-1.80) | † |
| | 5-26 weeks | 2.11 (1.75-2.54) | 1.10 (0.88-1.37) | † |
| | 27-52 weeks | 1.05 (0.83-1.33) | 0.88 (0.71-1.10) | † |
| | 1-2 years | 1.06 (0.90-1.25) | 0.89 (0.77-1.03) | † |
| | 2-3 years | 1.00 (0.85-1.18) | 0.83 (0.71-0.96) | † |
| | 3-4 years | 1.11 (0.95-1.29) | 0.81 (0.67-0.97) | † |
| | 4-5.5 years | 1.04 (0.85-1.28) | - | - |

† Insufficient events for estimation - Time period falls outside of cohort follow-up

**Supplementary Table 12: Maximally adjusted hazard ratios and 95% CIs for dementia following diagnosis of COVID-19 by cohort and by history of cognitive impairment**

| Outcome | Time since COVID-19 | Pre-vaccination cohort | Vaccinated cohort | Unvaccinated cohort |
|---|---|---|---|---|
| Prior history of cognitive impairment | 0-4 weeks | 3.58 (3.06-4.19) | 1.47 (1.15-1.88) | † |
| | 5-26 weeks | 1.30 (1.14-1.48) | 1.13 (0.99-1.30) | † |
| | 27-52 weeks | 1.03 (0.89-1.20) | 1.02 (0.88-1.20) | † |
| | 1-2 years | 1.17 (1.05-1.32) | 0.89 (0.78-1.02) | † |
| | 2-3 years | 1.18 (1.03-1.36) | 0.94 (0.81-1.10) | † |
| | 3-4 years | 1.20 (1.02-1.41) | 1.04 (0.86-1.27) | † |
| | 4-5.5 years | 1.08 (0.86-1.38) | - | - |
| No prior history of cognitive impairment | 0-4 weeks | 6.04 (5.43-6.72) | 2.57 (2.14-3.09) | 10.08 (6.65-15.28) |
| | 5-26 weeks | 2.40 (2.22-2.59) | 1.13 (1.01-1.26) | 1.70 (1.09-2.66) |
| | 27-52 weeks | 1.63 (1.50-1.77) | 0.95 (0.85-1.06) | 1.44 (0.91-2.27) |
| | 1-2 years | 1.21 (1.13-1.29) | 0.91 (0.84-0.98) | 1.22 (0.85-1.75) |
| | 2-3 years | 1.13 (1.05-1.21) | 0.89 (0.83-0.95) | 1.12 (0.76-1.65) |
| | 3-4 years | 1.15 (1.07-1.23) | 0.85 (0.78-0.93) | 1.42 (0.95-2.14) |
| | 4-5.5 years | 1.09 (0.99-1.20) | - | - |

† Insufficient events for estimation — - Time period falls outside of cohort follow-up

**Supplementary Table 13: Maximally adjusted hazard ratios and 95% CIs for dementia following diagnosis of COVID-19 by cohort and by history of Parkinson's disease**

| Subgroup | Time since COVID-19 | Pre-vaccination cohort | Vaccinated cohort | Unvaccinated cohort |
|---|---|---|---|---|
| **Prior history of Parkinson's disease** | 0-4 weeks | 5.12 (3.62-7.25) | | † |
| | 5-26 weeks | 1.45 (1.06-1.98) | 1.08 (0.81-1.43) | † |
| | 27-52 weeks | 1.36 (0.98-1.87) | | † |
| | 1-2 years | 1.09 (0.82-1.45) | 1.17 (0.86-1.59) | † |
| | 2-3 years | 1.24 (0.90-1.71) | 1.10 (0.84-1.44) | † |
| | 3-4 years | 1.78 (1.30-2.42) | | † |
| | 4-5.5 years | 1.33 (0.83-2.14) | - | - |
| **No prior history of Parkinson's disease** | 0-4 weeks | 5.05 (4.61-5.54) | 2.03 (1.74-2.36) | 8.60 (6.03-12.26) |
| | 5-26 weeks | 2.04 (1.90-2.18) | 1.15 (1.05-1.25) | 1.42 (0.95-2.11) |
| | 27-52 weeks | 1.44 (1.34-1.55) | 0.98 (0.89-1.07) | 1.40 (0.94-2.07) |
| | 1-2 years | 1.19 (1.12-1.26) | 0.89 (0.83-0.95) | 1.14 (0.82-1.59) |
| | 2-3 years | 1.12 (1.05-1.19) | 0.89 (0.84-0.95) | 1.04 (0.72-1.49) |
| | 3-4 years | 1.12 (1.05-1.19) | 0.84 (0.78-0.91) | 1.44 (0.99-2.12) |
| | 4-5.5 years | 1.06 (0.97-1.16) | - | - |

† Insufficient events for estimation - Time period falls outside of cohort follow-up

**Supplementary Table 14: Maximally adjusted hazard ratios and 95% confidence intervals for common dementia subtypes following diagnosis of COVID-19 by cohort, overall, and by COVID-19 severity**

| Outcome | Time since COVID-19 | Pre-vaccination cohort | Vaccinated cohort | Unvaccinated cohort |
|---|---|---|---|---|
| **All COVID-19** | | | | |
| | 0-4 weeks | 2.29 (1.85-2.82) | 0.84 (0.60-1.18) | |
| | 5-26 weeks | 1.53 (1.35-1.72) | 1.15 (1.01-1.30) | 1.55 (1.01-2.39) |
| | 27-52 weeks | 1.47 (1.32-1.64) | 0.93 (0.82-1.06) | |
| **Alzheimer's disease** | 1-2 years | 1.06 (0.97-1.16) | 0.90 (0.82-0.98) | 1.04 (0.62-1.77) |
| | 2-3 years | 0.97 (0.89-1.06) | 0.89 (0.82-0.96) | 0.95 (0.61-1.49) |
| | 3-4 years | 1.04 (0.95-1.14) | 0.88 (0.80-0.98) | |
| | 4-5.5 years | 0.98 (0.87-1.10) | - | - |
| | 0-4 weeks | 4.34 (3.51-5.37) | 1.76 (1.20-2.56) | † |
| | 5-26 weeks | 2.53 (2.22-2.88) | 1.03 (0.83-1.29) | † |
| | 27-52 weeks | 1.71 (1.47-1.98) | 1.04 (0.85-1.27) | † |
| **Vascular dementia** | 1-2 years | 1.46 (1.30-1.64) | 1.05 (0.92-1.21) | † |
| | 2-3 years | 1.35 (1.19-1.53) | 0.98 (0.85-1.13) | † |
| | 3-4 years | 1.26 (1.10-1.44) | 0.87 (0.73-1.06) | † |
| | 4-5.5 years | 1.14 (0.95-1.37) | - | - |
| | 0-1 years | 1.55 (1.10-2.19) | 1.23 (0.84-1.79) | † |
| **Lewy body dementia** | 1-2 years | 1.01 (0.65-1.55) | 0.91 (0.60-1.39) | † |
| | 2-4 years | 0.73 (0.51-1.05) | 1.03 (0.77-1.39) | † |
| | 4-5.5 years | 0.85 (0.47-1.55) | - | - |
| **Hospitalised COVID-19** | | | | |
| | 0-4 weeks | 4.21 (1.85-9.53) | | † |
| | 5-26 weeks | 1.33 (0.70-2.51) | 2.07 (1.70-2.52) | † |
| | 27-52 weeks | 1.56 (0.90-2.69) | | † |
| **Alzheimer's disease** | 1-2 years | 1.21 (0.77-1.89) | 1.30 (1.00-1.69) | † |
| | 2-3 years | 1.03 (0.62-1.71) | 1.22 (0.98-1.53) | † |
| | 3-4 years | 0.58 (0.26-1.30) | | † |
| | 4-5.5 years | - | - | - |
| | 0-4 weeks | 4.21 (1.85-9.53) | | † |
| | 5-26 weeks | 1.33 (0.70-2.51) | 2.43 (1.85-3.19) | † |
| | 27-52 weeks | 1.56 (0.90-2.69) | | † |
| **Vascular dementia** | 1-2 years | 1.21 (0.77-1.89) | 1.43 (0.98-2.09) | † |
| | 2-3 years | 1.03 (0.62-1.71) | 1.46 (1.06-2.02) | † |
| | 3-4 years | 0.58 (0.26-1.30) | | † |
| | 4-5.5 years | - | - | - |
| **Non-hospitalised COVID-19** | | | | |
| | 0-4 weeks | 1.54 (1.15-2.06) | 0.71 (0.48-1.05) | † |
| | 5-26 weeks | 1.38 (1.20-1.58) | 1.05 (0.91-1.20) | † |
| | 27-52 weeks | 1.30 (1.14-1.48) | 0.83 (0.72-0.95) | † |
| **Alzheimer's disease** | 1-2 years | 0.96 (0.86-1.06) | 0.86 (0.79-0.94) | † |
| | 2-3 years | 0.93 (0.84-1.03) | 0.85 (0.78-0.93) | † |
| | 3-4 years | 1.02 (0.93-1.12) | 0.87 (0.79-0.97) | † |
| | 4-5.5 years | 0.94 (0.82-1.07) | - | - |
| | 0-4 weeks | 2.38 (1.72-3.31) | 1.15 (0.69-1.90) | † |
| | 5-26 weeks | 2.28 (1.95-2.66) | 0.90 (0.70-1.16) | † |
| | 27-52 weeks | 1.44 (1.20-1.72) | 0.90 (0.71-1.13) | † |
| **Vascular dementia** | 1-2 years | 1.28 (1.11-1.47) | 1.01 (0.87-1.17) | † |
| | 2-3 years | 1.23 (1.06-1.42) | 0.94 (0.81-1.10) | † |
| | 3-4 years | 1.24 (1.07-1.44) | 0.82 (0.67-1.00) | † |
| | 4-5.5 years | 1.08 (0.87-1.34) | - | - |
| | 0-1 years | 1.27 (0.83-1.94) | 1.14 (0.76-1.72) | † |
| **Lewy body dementia** | 1-2 years | 0.83 (0.49-1.41) | 0.89 (0.57-1.38) | † |
| | 2-4 years | 0.79 (0.54-1.15) | 1.07 (0.79-1.44) | † |
| | 4-5.5 years | 0.76 (0.38-1.53) | - | - |

† Insufficient events for estimation — - Time period falls outside of cohort follow-up

**Supplementary Table 15: Maximally adjusted hazard ratios and 95% confidence intervals for Parkinson's disease and associated risk conditions following diagnosis of COVID-19 by cohort, overall, and by COVID-19 severity**

| Outcome | Time since COVID-19 | Pre-vaccination cohort | Vaccinated cohort | Unvaccinated cohort |
|---|---|---|---|---|
| **All COVID-19** | | | | |
| **Parkinson's disease** | 0-4 weeks | 4.03 (3.07-5.29) | 1.67 (1.18-2.36) | † |
| | 5-26 weeks | 1.23 (1.00-1.52) | 0.96 (0.79-1.18) | † |
| | 27-52 weeks | 1.08 (0.89-1.31) | 0.69 (0.56-0.86) | † |
| | 1-2 years | 0.99 (0.86-1.14) | 0.96 (0.84-1.09) | † |
| | 2-3 years | 1.02 (0.89-1.17) | 0.91 (0.79-1.04) | † |
| | 3-4 years | 0.93 (0.81-1.08) | 0.87 (0.74-1.03) | † |
| | 4-5.5 years | 1.09 (0.90-1.31) | - | - |
| **Restless leg syndrome** | 0-4 weeks | 0.90 (0.61-1.35) | 0.82 (0.54-1.26) | |
| | 5-26 weeks | 1.34 (1.16-1.55) | 1.09 (0.92-1.28) | 1.39 (0.99-1.95) |
| | 27-52 weeks | 1.26 (1.09-1.45) | 1.05 (0.90-1.22) | |
| | 1-2 years | 1.20 (1.08-1.34) | 1.16 (1.05-1.29) | 1.95 (1.46-2.61) |
| | 2-3 years | 1.09 (0.98-1.22) | 1.16 (1.05-1.28) | 1.27 (0.98-1.64) |
| | 3-4 years | 1.29 (1.17-1.42) | 1.14 (1.00-1.29) | |
| | 4-5.5 years | 1.13 (0.98-1.31) | - | - |
| **Rapid eye movement sleep disorder** | 0-4 weeks | 8.80 (8.14-9.50) | 2.86 (2.54-3.21) | 10.78 (8.52-13.63) |
| | 5-26 weeks | 1.27 (1.18-1.38) | 1.16 (1.07-1.26) | 1.56 (1.20-2.03) |
| | 27-52 weeks | 1.31 (1.22-1.41) | 1.05 (0.97-1.13) | 1.32 (1.03-1.70) |
| | 1-2 years | 1.18 (1.12-1.24) | 1.14 (1.08-1.19) | 1.37 (1.16-1.61) |
| | 2-3 years | 1.26 (1.20-1.32) | 1.18 (1.13-1.23) | 1.47 (1.27-1.71) |
| | 3-4 years | 1.31 (1.25-1.37) | 1.19 (1.13-1.26) | 1.47 (1.24-1.75) |
| | 4-5.5 years | 1.33 (1.25-1.41) | - | - |
| **Hospitalised COVID-19** | | | | |
| **Parkinson's disease** | 0-4 weeks | 15.13 (11.46-19.96) | | † |
| | 5-26 weeks | 1.29 (0.77-2.19) | 1.79 (1.20-2.68) | † |
| | 27-52 weeks | 1.68 (1.13-2.51) | | † |
| | 1-2 years | 1.25 (0.90-1.74) | 1.27 (0.77-2.11) | † |
| | 2-3 years | 1.28 (0.92-1.79) | 1.11 (0.70-1.73) | † |
| | 3-4 years | 1.45 (1.05-2.01) | | † |
| | 4-5.5 years | 1.52 (1.02-2.27) | - | - |
| **Restless leg syndrome** | 0-1 years | 1.34 (0.94-1.89) | † | † |
| | 1-2 years | 1.31 (0.89-1.92) | † | † |
| | 2-4 years | 1.53 (1.18-1.98) | † | † |
| | 4-5.5 years | 1.07 (0.62-1.84) | - | - |
| **Rapid eye movement sleep disorder** | 0-4 weeks | 70.90 (64.84-77.52) | 50.49 (43.69-58.34) | |
| | 5-26 weeks | 2.00 (1.63-2.45) | 2.59 (1.93-3.47) | 9.24 (7.38-11.57) |
| | 27-52 weeks | 1.96 (1.62-2.38) | 1.48 (1.04-2.11) | |
| | 1-2 years | 1.94 (1.69-2.23) | 1.57 (1.23-2.00) | 2.78 (1.94-3.98) |
| | 2-3 years | 1.77 (1.53-2.04) | 1.99 (1.60-2.46) | 2.22 (1.67-2.96) |
| | 3-4 years | 2.06 (1.81-2.35) | 1.55 (1.16-2.09) | |
| | 4-5.5 years | 1.85 (1.57-2.19) | - | - |
| **Non-hospitalised COVID-19** | | | | |
| **Parkinson's disease** | 0-4 weeks | 2.52 (1.73-3.68) | 1.24 (0.81-1.88) | † |
| | 5-26 weeks | 1.22 (0.97-1.54) | 0.95 (0.77-1.17) | † |
| | 27-52 weeks | 0.98 (0.79-1.22) | 0.67 (0.53-0.84) | † |
| | 1-2 years | 0.95 (0.81-1.11) | 0.94 (0.82-1.07) | † |
| | 2-3 years | 0.98 (0.84-1.14) | 0.89 (0.77-1.02) | † |
| | 3-4 years | 0.85 (0.72-1.01) | 0.88 (0.74-1.04) | † |
| | 4-5.5 years | 1.01 (0.82-1.24) | - | - |
| **Restless leg syndrome** | 0-4 weeks | 0.94 (0.62-1.42) | 0.77 (0.49-1.20) | |
| | 5-26 weeks | 1.33 (1.15-1.55) | 1.07 (0.90-1.26) | 1.30 (0.91-1.86) |
| | 27-52 weeks | 1.23 (1.06-1.42) | 1.01 (0.87-1.19) | |

| | | | | |
|---|---|---|---|---|
| | 1-2 years | 1.19 (1.06-1.33) | 1.15 (1.04-1.28) | 1.87 (1.38-2.54) |
| | 2-3 years | 1.06 (0.94-1.19) | 1.15 (1.04-1.27) | 1.18 (0.90-1.55) |
| | 3-4 years | 1.26 (1.14-1.39) | 1.15 (1.01-1.30) | |
| | 4-5.5 years | 1.13 (0.97-1.31) | - | - |
| **Rapid eye movement sleep disorder** | 0-4 weeks | 1.63 (1.36-1.96) | 0.96 (0.79-1.18) | 1.33 (1.09-1.61) |
| | 5-26 weeks | 1.19 (1.10-1.30) | 1.12 (1.03-1.21) | |
| | 27-52 weeks | 1.24 (1.15-1.34) | 1.03 (0.96-1.11) | |
| | 1-2 years | 1.10 (1.03-1.16) | 1.12 (1.07-1.18) | 1.21 (1.00-1.45) |
| | 2-3 years | 1.21 (1.15-1.28) | 1.16 (1.10-1.21) | 1.38 (1.22-1.57) |
| | 3-4 years | 1.24 (1.18-1.30) | 1.18 (1.12-1.25) | |
| | 4-5.5 years | 1.27 (1.19-1.35) | - | - |

† Insufficient events for estimation
- Time period falls outside of cohort follow-up

**Supplementary Table 16: Maximally adjusted hazard ratios and 95% CIs for Parkinson's disease following diagnosis of COVID-19 by cohort and by history of Parkinson's risk conditions (defined as restless leg syndrome and/or REM sleep disorder**

| Outcome | Time since COVID-19 | Pre-vaccination cohort | Vaccinated cohort | Unvaccinated cohort |
|---|---|---|---|---|
| **Prior history of Parkinson's risk conditions** | 0-1 years | 1.96 (1.23-3.14) | † | † |
| | 1-2 years | 1.52 (0.87-2.65) | † | † |
| | 2-4 years | 1.32 (0.86-2.02) | † | † |
| | 4-5.5 years | 0.45 (0.11-1.80) | - | - |
| **No prior history of Parkinson's risk conditions** | 0-4 weeks | 3.97 (2.99-5.27) | 1.60 (1.11-2.31) | † |
| | 5-26 weeks | 1.23 (0.99-1.53) | 0.97 (0.79-1.19) | † |
| | 27-52 weeks | 1.04 (0.85-1.27) | 0.71 (0.57-0.88) | † |
| | 1-2 years | 0.97 (0.84-1.12) | 0.94 (0.82-1.08) | † |
| | 2-3 years | 0.99 (0.86-1.15) | 0.91 (0.79-1.04) | † |
| | 3-4 years | 0.94 (0.81-1.10) | 0.87 (0.73-1.03) | † |
| | 4-5.5 years | 1.12 (0.93-1.35) | - | - |

† Insufficient events for estimation - Time period falls outside of cohort follow-up

**Supplementary Table 17: Maximally adjusted hazard ratios and 95% confidence intervals for other neurogenerative and neurological conditions following diagnosis of COVID-19 by cohort, overall, and by COVID-19 severity**

| Outcome | Time since COVID-19 | Pre-vaccination cohort | Vaccinated cohort | Unvaccinated cohort |
|---|---|---|---|---|
| **All COVID-19** | | | | |
| **Migraine** | 0-4 weeks | 1.33 (1.20-1.48) | 1.15 (1.02-1.30) | 1.78 (1.39-2.26) |
| | 5-26 weeks | 1.29 (1.24-1.35) | 1.14 (1.08-1.20) | 1.32 (1.16-1.49) |
| | 27-52 weeks | 1.21 (1.15-1.26) | 1.14 (1.09-1.20) | 1.58 (1.42-1.76) |
| | 1-2 years | 1.21 (1.17-1.25) | 1.11 (1.07-1.15) | 1.48 (1.36-1.60) |
| | 2-3 years | 1.19 (1.15-1.23) | 1.10 (1.06-1.14) | 1.38 (1.27-1.51) |
| | 3-4 years | 1.15 (1.11-1.19) | 1.13 (1.07-1.18) | 1.19 (1.06-1.34) |
| | 4-5.5 years | 1.19 (1.13-1.25) | - | - |
| **Motor neurone disease** | 0-1 years | 1.11 (0.80-1.53) | 0.70 (0.48-1.03) | † |
| | 1-2 years | 0.90 (0.62-1.31) | 0.76 (0.53-1.09) | † |
| | 2-4 years | 1.09 (0.86-1.39) | 1.03 (0.80-1.32) | † |
| | 4-5.5 years | 1.27 (0.82-1.96) | - | - |
| **Multiple sclerosis** | 0-4 weeks | 2.42 (1.65-3.54) | | |
| | 5-26 weeks | 1.10 (0.86-1.39) | 0.95 (0.79-1.13) | 1.68 (1.15-2.46) |
| | 27-52 weeks | 0.93 (0.73-1.19) | | |
| | 1-2 years | 0.82 (0.68-0.99) | 0.94 (0.78-1.12) | 1.21 (0.77-1.89) |
| | 2-3 years | 0.85 (0.70-1.04) | 0.98 (0.85-1.14) | 0.85 (0.55-1.31) |
| | 3-4 years | 1.00 (0.83-1.21) | | |
| | 4-5.5 years | 0.88 (0.67-1.17) | - | - |
| **Hospitalised COVID-19** | | | | |
| **Migraine** | 0-4 weeks | 8.42 (6.61-10.72) | | |
| | 5-26 weeks | 1.08 (0.81-1.44) | 1.95 (1.47-2.58) | 2.19 (1.59-3.02) |
| | 27-52 weeks | 1.31 (1.02-1.70) | | |
| | 1-2 years | 1.17 (0.95-1.44) | 1.43 (1.01-2.02) | 1.19 (0.77-1.85) |
| | 2-3 years | 1.04 (0.83-1.30) | 1.42 (1.07-1.89) | 1.33 (0.95-1.86) |
| | 3-4 years | 1.12 (0.90-1.41) | | |
| | 4-5.5 years | 1.42 (1.09-1.85) | - | - |
| **Non-hospitalised COVID-19** | | | | |
| **Migraine** | 0-4 weeks | 1.12 (1.00-1.25) | 1.09 (0.96-1.23) | 1.44 (1.10-1.90) |
| | 5-26 weeks | 1.29 (1.24-1.35) | 1.14 (1.08-1.20) | 1.32 (1.16-1.50) |
| | 27-52 weeks | 1.20 (1.15-1.26) | 1.14 (1.08-1.20) | 1.59 (1.42-1.77) |
| | 1-2 years | 1.21 (1.17-1.25) | 1.11 (1.07-1.15) | 1.49 (1.38-1.62) |
| | 2-3 years | 1.19 (1.15-1.23) | 1.10 (1.06-1.14) | 1.39 (1.27-1.52) |
| | 3-4 years | 1.15 (1.11-1.19) | 1.12 (1.06-1.17) | 1.19 (1.05-1.34) |
| | 4-5.5 years | 1.18 (1.12-1.24) | - | - |
| **Motor neurone disease** | 0-1 years | 1.01 (0.70-1.44) | 0.62 (0.41-0.94) | † |
| | 1-2 years | 0.89 (0.60-1.33) | 0.73 (0.50-1.06) | † |
| | 2-4 years | 1.03 (0.79-1.34) | 1.02 (0.79-1.31) | † |
| | 4-5.5 years | 1.29 (0.82-2.04) | - | - |
| **Multiple sclerosis** | 0-4 weeks | 1.63 (1.01-2.63) | | |
| | 5-26 weeks | 1.01 (0.78-1.30) | 0.91 (0.76-1.09) | 1.43 (0.94-2.17) |
| | 27-52 weeks | 0.91 (0.70-1.17) | | |
| | 1-2 years | 0.80 (0.66-0.98) | 0.94 (0.78-1.12) | 1.22 (0.77-1.93) |
| | 2-3 years | 0.85 (0.69-1.04) | 0.98 (0.84-1.13) | 0.78 (0.49-1.24) |
| | 3-4 years | 1.01 (0.84-1.23) | | |
| | 4-5.5 years | 0.88 (0.66-1.17) | - | - |

† Insufficient events for estimation  - Time period falls outside of cohort follow-up

**Supplementary Figure 1: Diagram showing cohort construction**

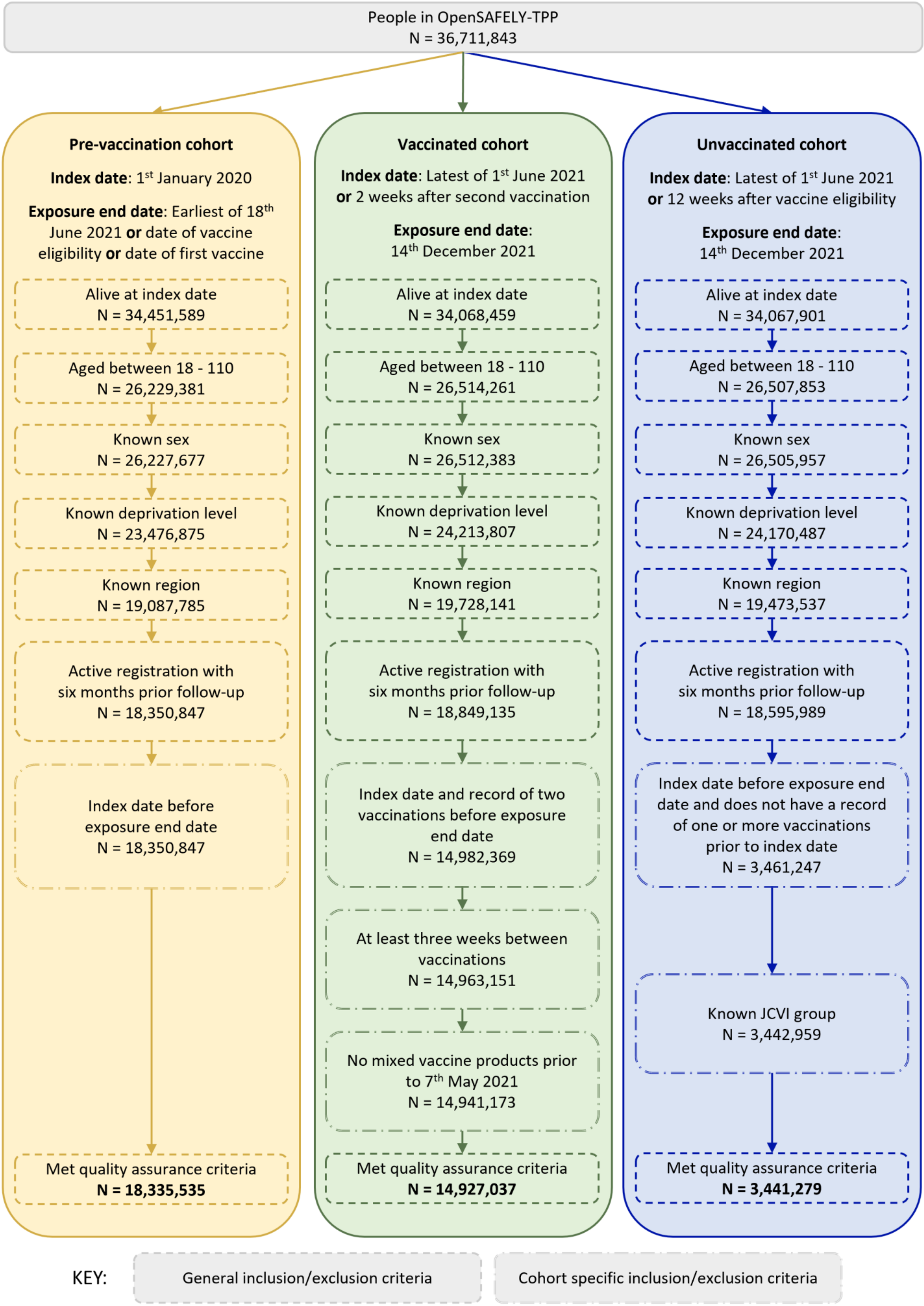

**Supplementary Figure 2: Maximally adjusted hazard ratios and 95% CIs for cognitive impairment and dementia following diagnosis of COVID-19 by cohort in people with a prior history of COVID-19**

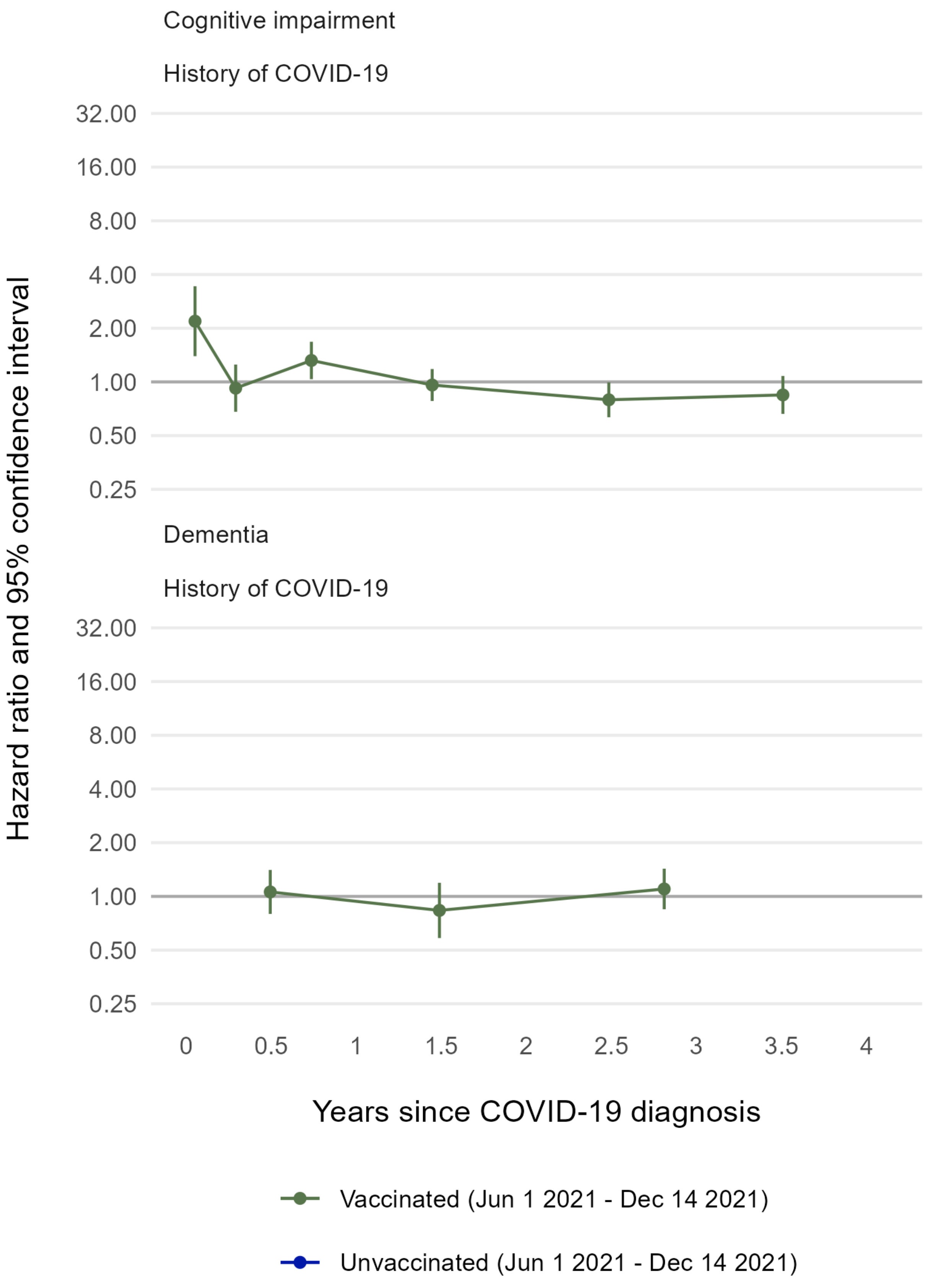

**Supplementary Figure 3: Maximally adjusted hazard ratios and 95% CIs for cognitive impairment and dementia following diagnosis of COVID-19 by cohort and by age group in years**

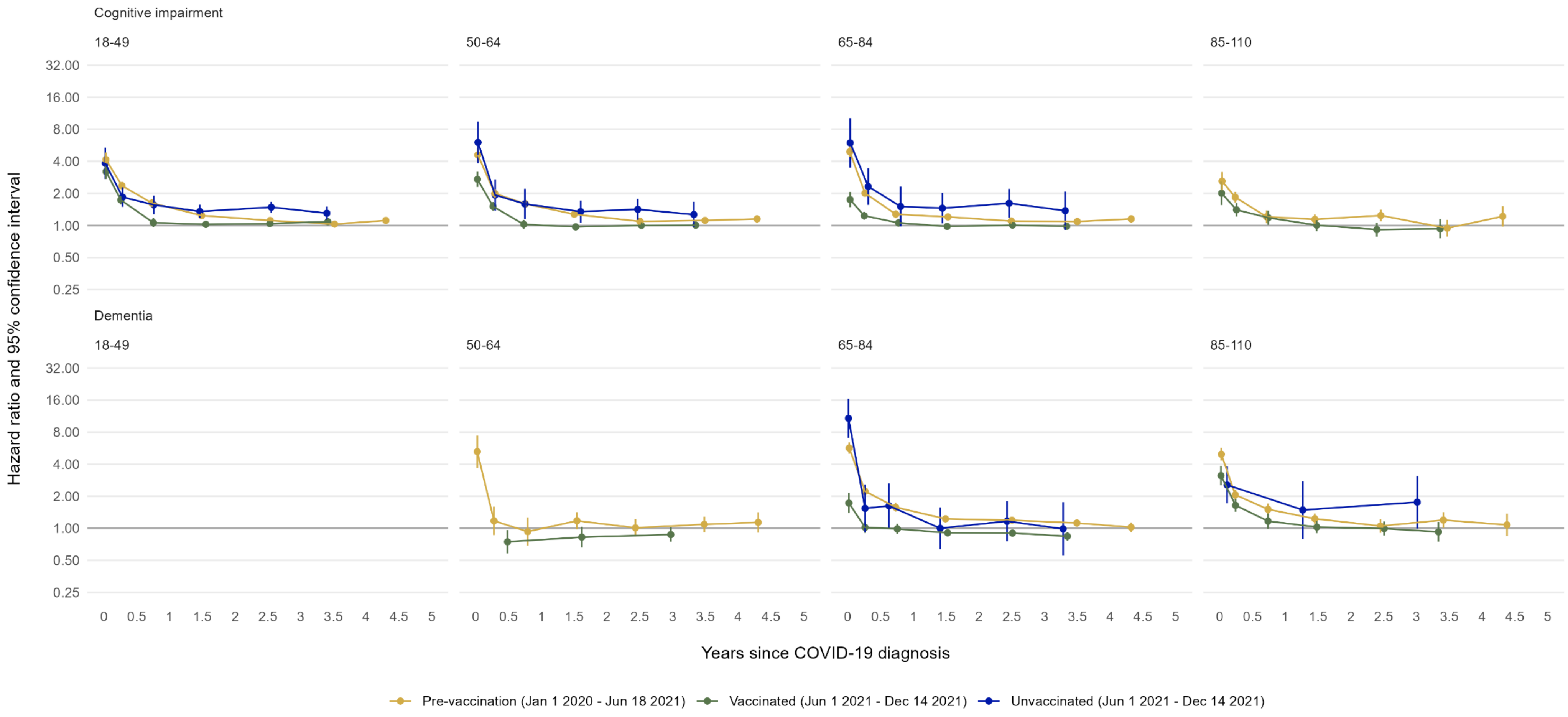

The age group 18-49 was excluded for the outcome dementia

**Supplementary Figure 4: Maximally adjusted hazard ratios and 95% CIs for cognitive impairment and dementia following diagnosis of COVID-19 by cohort and by sex**

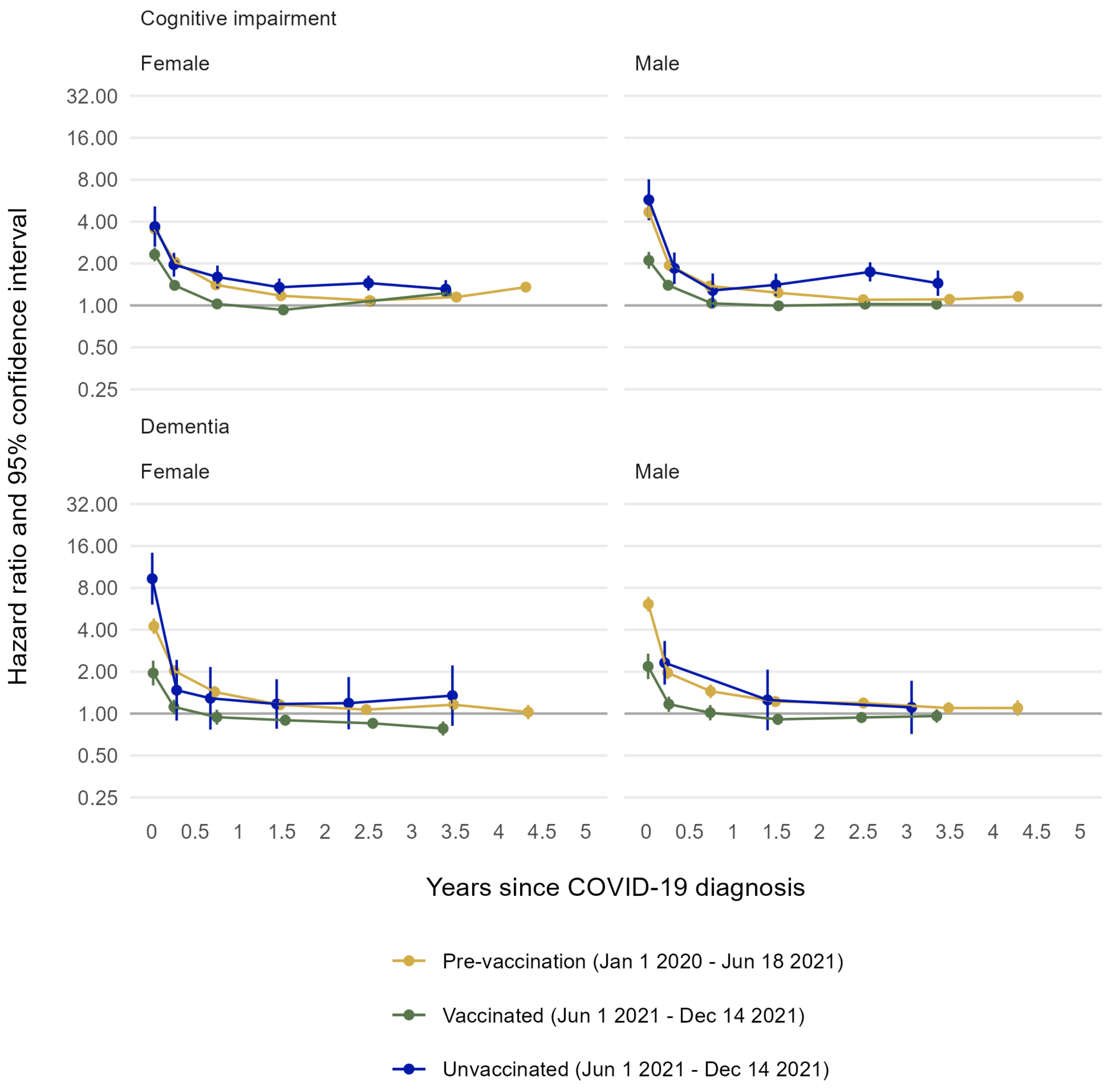

**Supplementary Figure 5: Maximally adjusted hazard ratios and 95% CIs for cognitive impairment and dementia following diagnosis of COVID-19 by cohort and by ethnicity**

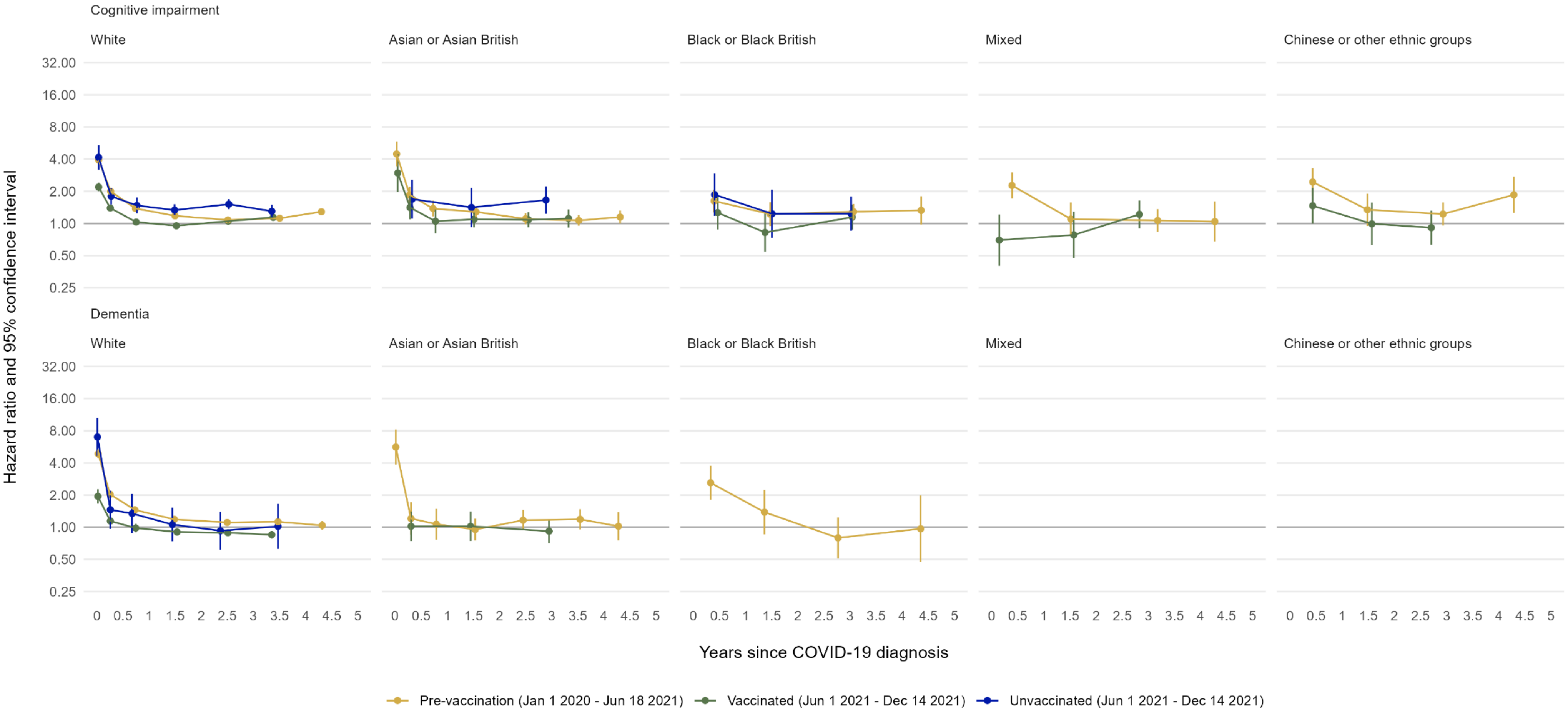

**Supplementary Figure 6: Maximally adjusted hazard ratios and 95% CIs for cognitive impairment and dementia following diagnosis of COVID-19 by cohort and by vascular risk (defined as history of hypertension and/or diabetes)**

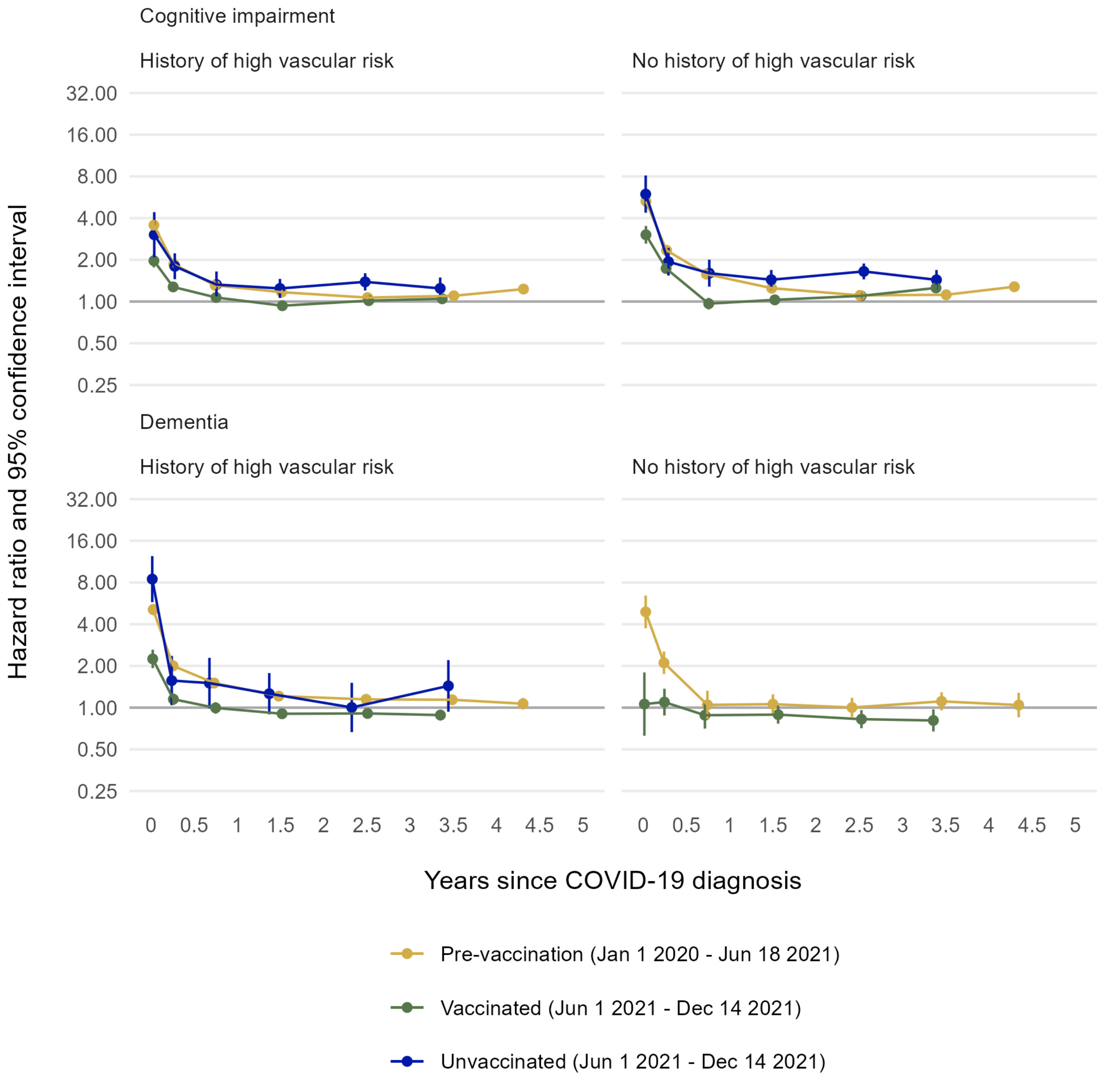

**Supplementary Figure 7: Maximally adjusted hazard ratios and 95% CIs for dementia following diagnosis of COVID-19 by cohort and by prior history of cognitive impairment**

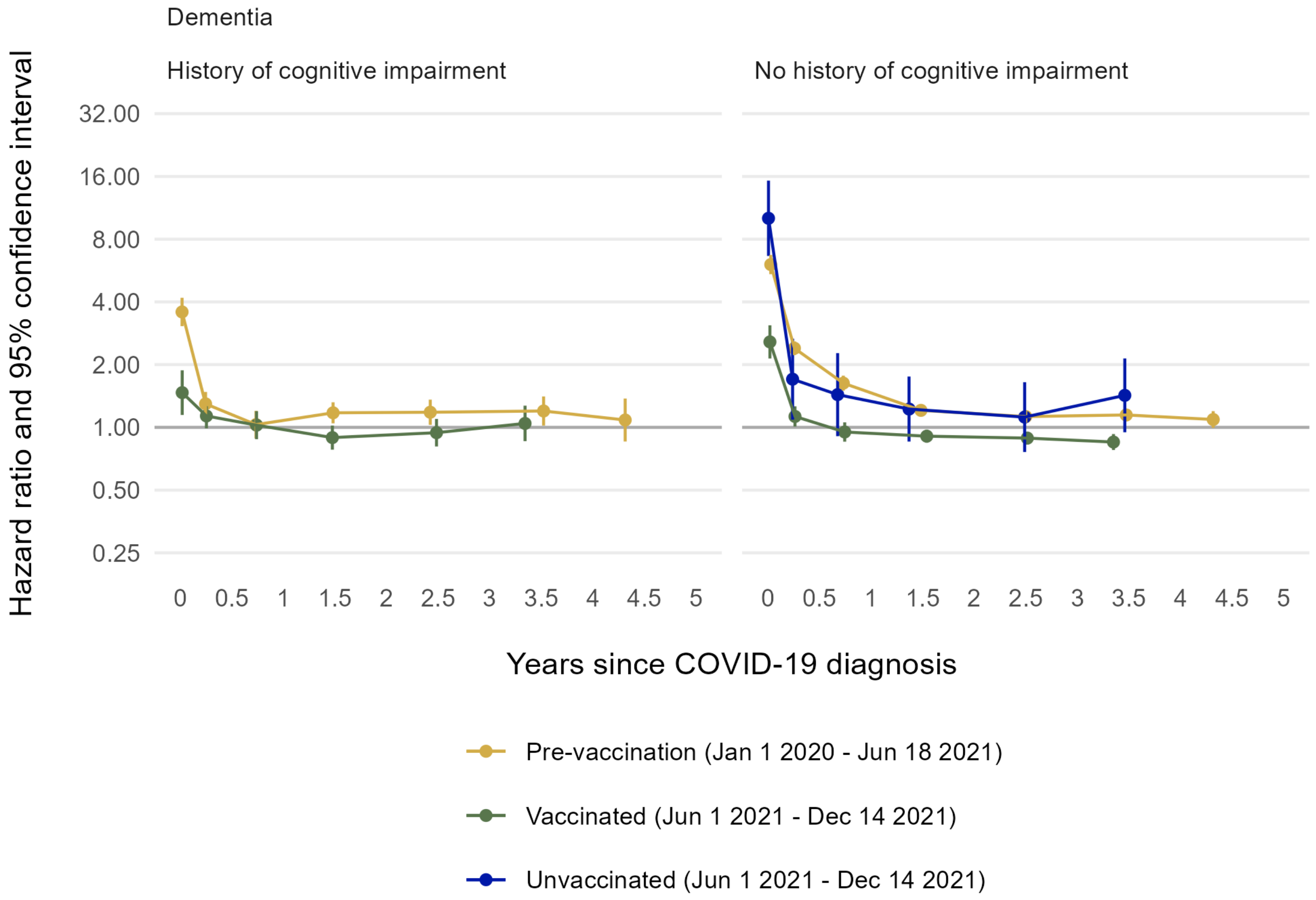

**Supplementary Figure 8: Maximally adjusted hazard ratios and 95% CIs for dementia following diagnosis of COVID-19 by cohort and by prior history of Parkinson's disease**

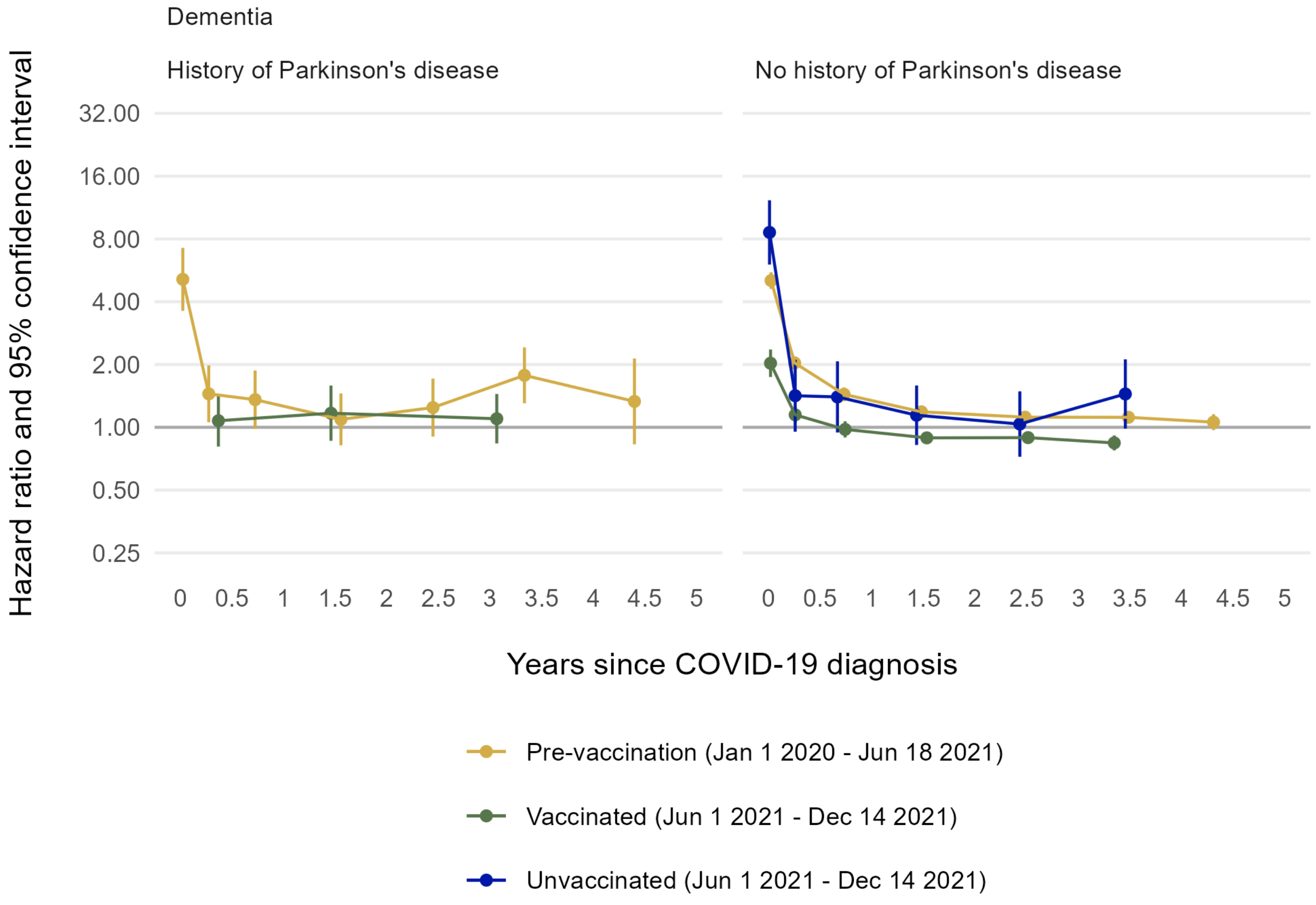

**Supplementary Figure 9: Maximally adjusted hazard ratios and 95% CIs for common dementia subtypes following diagnosis of COVID-19 by cohort, overall, and by COVID-19 severity**

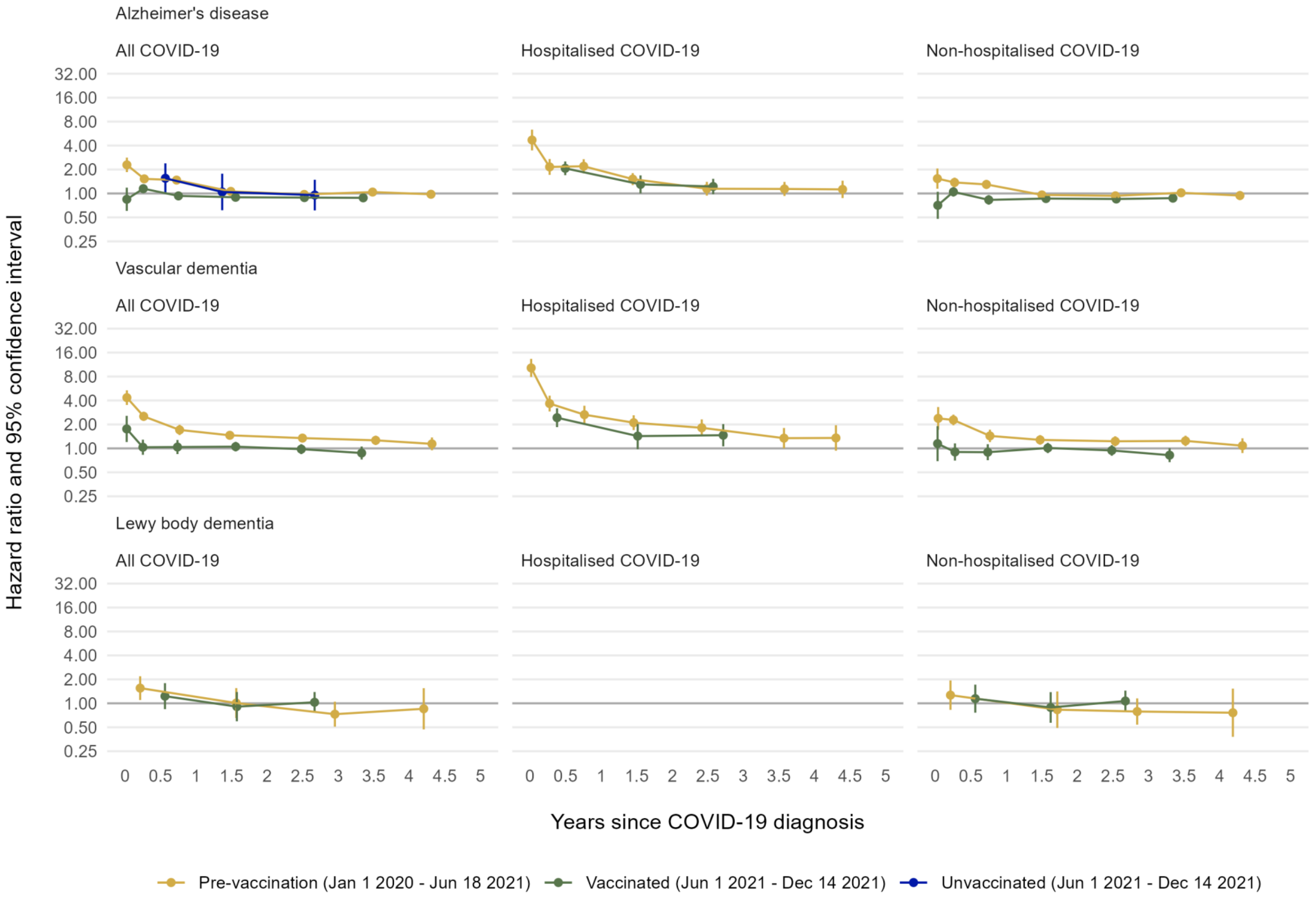

**Supplementary Figure 10 Maximally adjusted hazard ratios and 95% CIs for Parkinson's disease and associated risk conditions following diagnosis of COVID-19 by cohort, overall, and by COVID-19 severity**

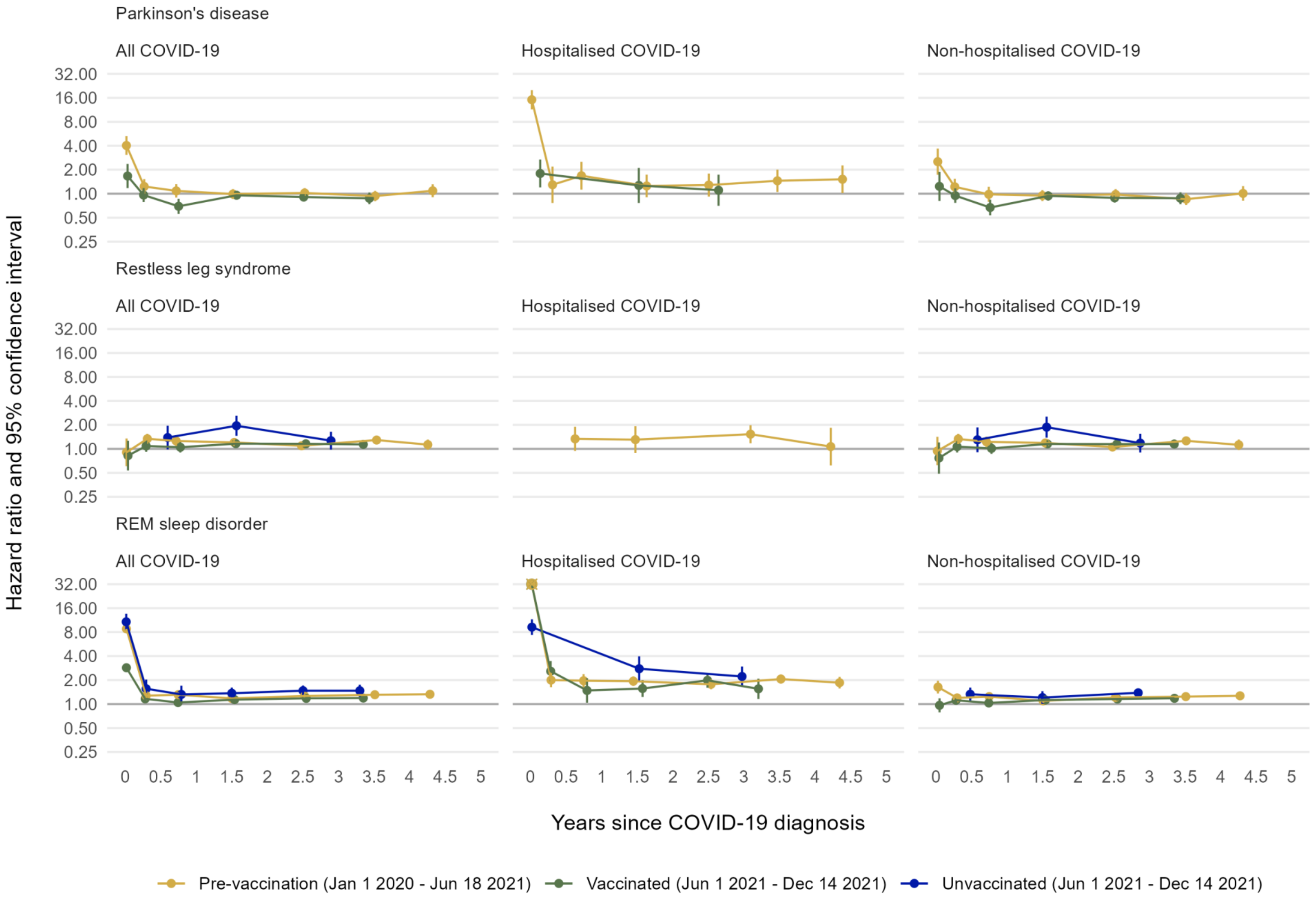

**Supplementary Figure 11: Maximally adjusted hazard ratios and 95% CIs for Parkinson's disease following diagnosis of COVID-19 by cohort and by prior history of Parkinson's risk conditions (defined as restless leg syndrome and/or REM sleep disorder)**

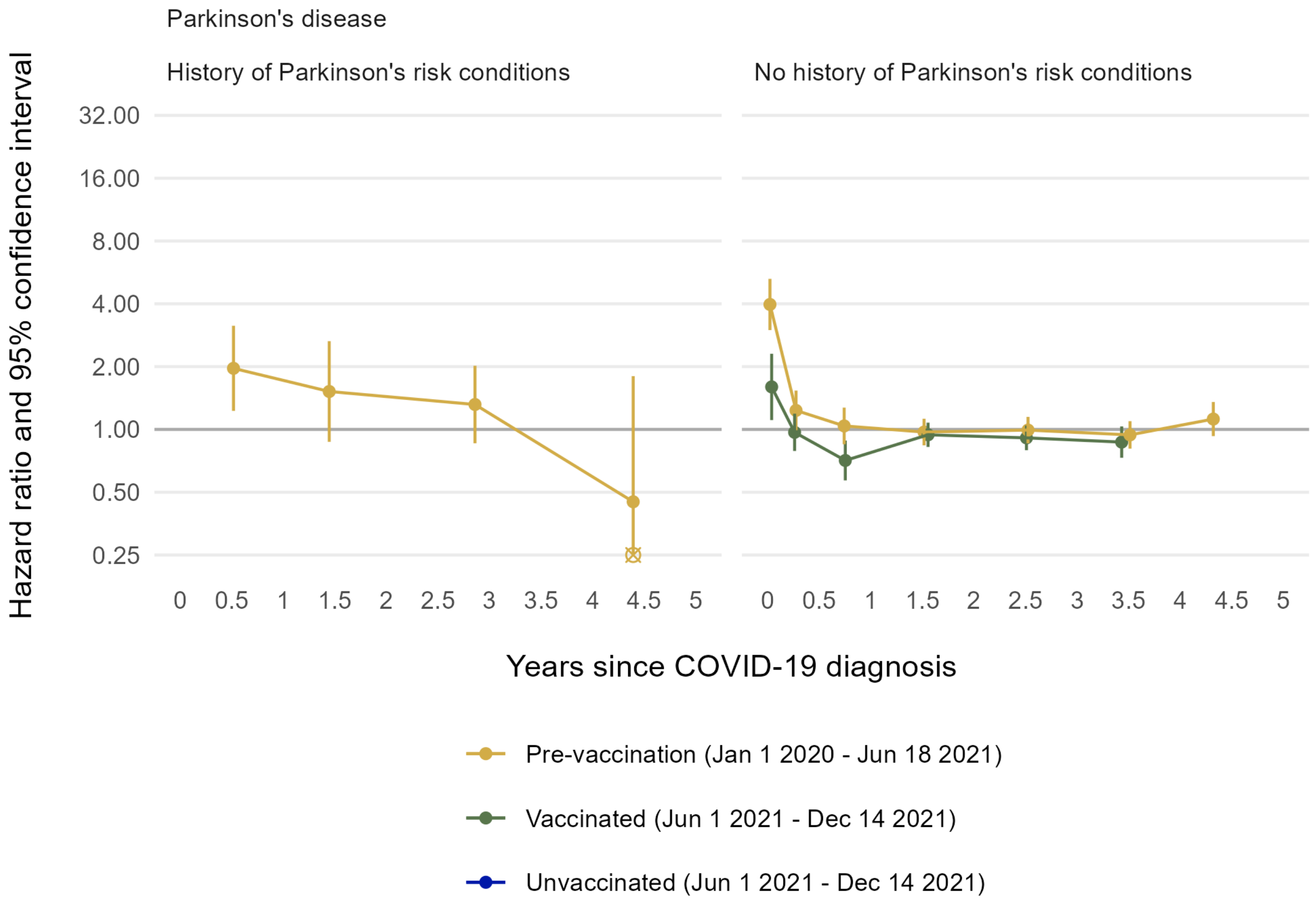

**Supplementary Figure 12: Maximally adjusted hazard ratios and 95% CIs for other neurogenerative and neurological conditions following diagnosis of COVID-19 by cohort, overall, and by COVID-19 severity**

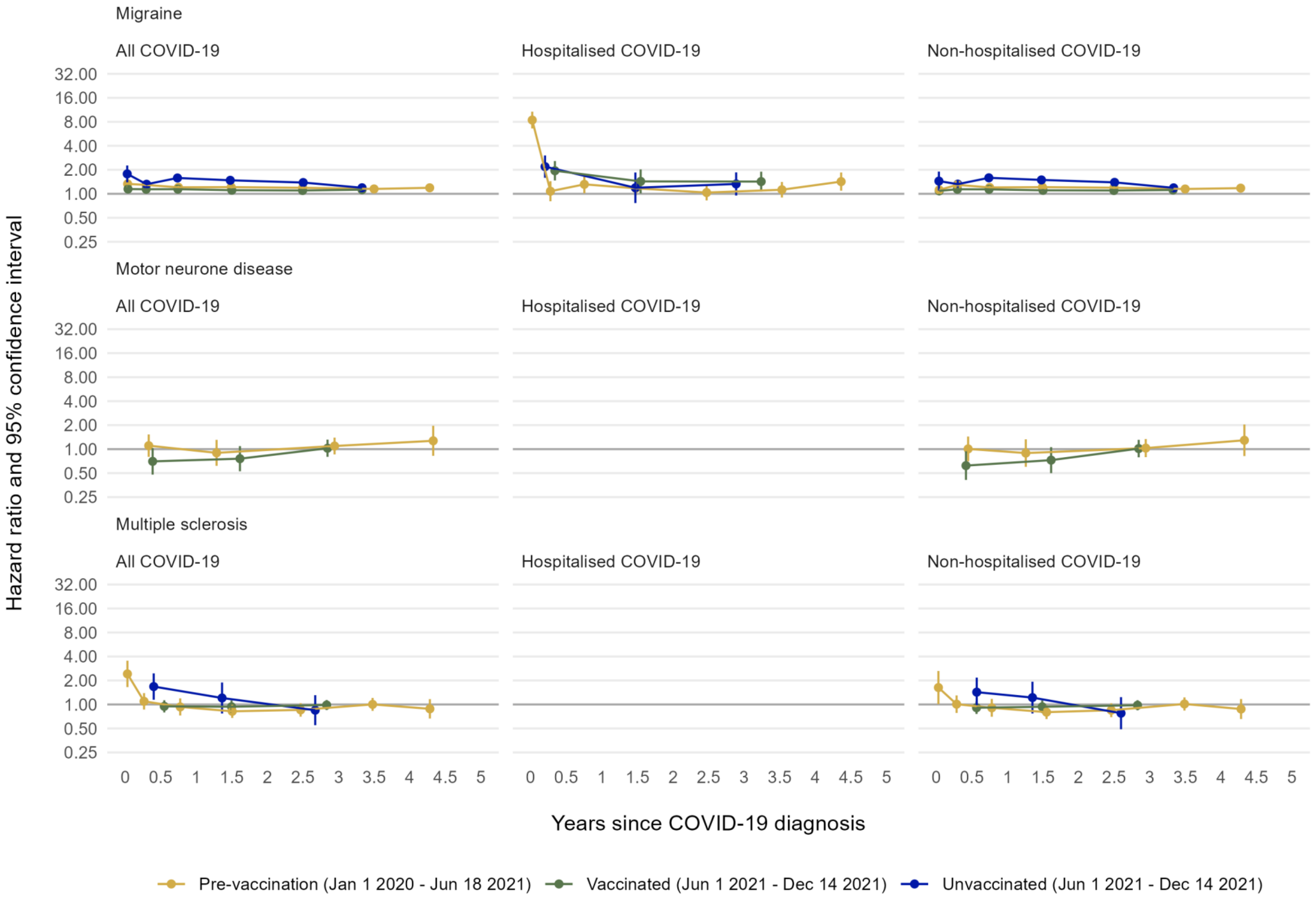

**Supplementary Figure 13: Absolute excess risk up to 52 weeks for cognitive impairment and dementia following diagnosis of COVID-19 by cohort (starting from day 1)**

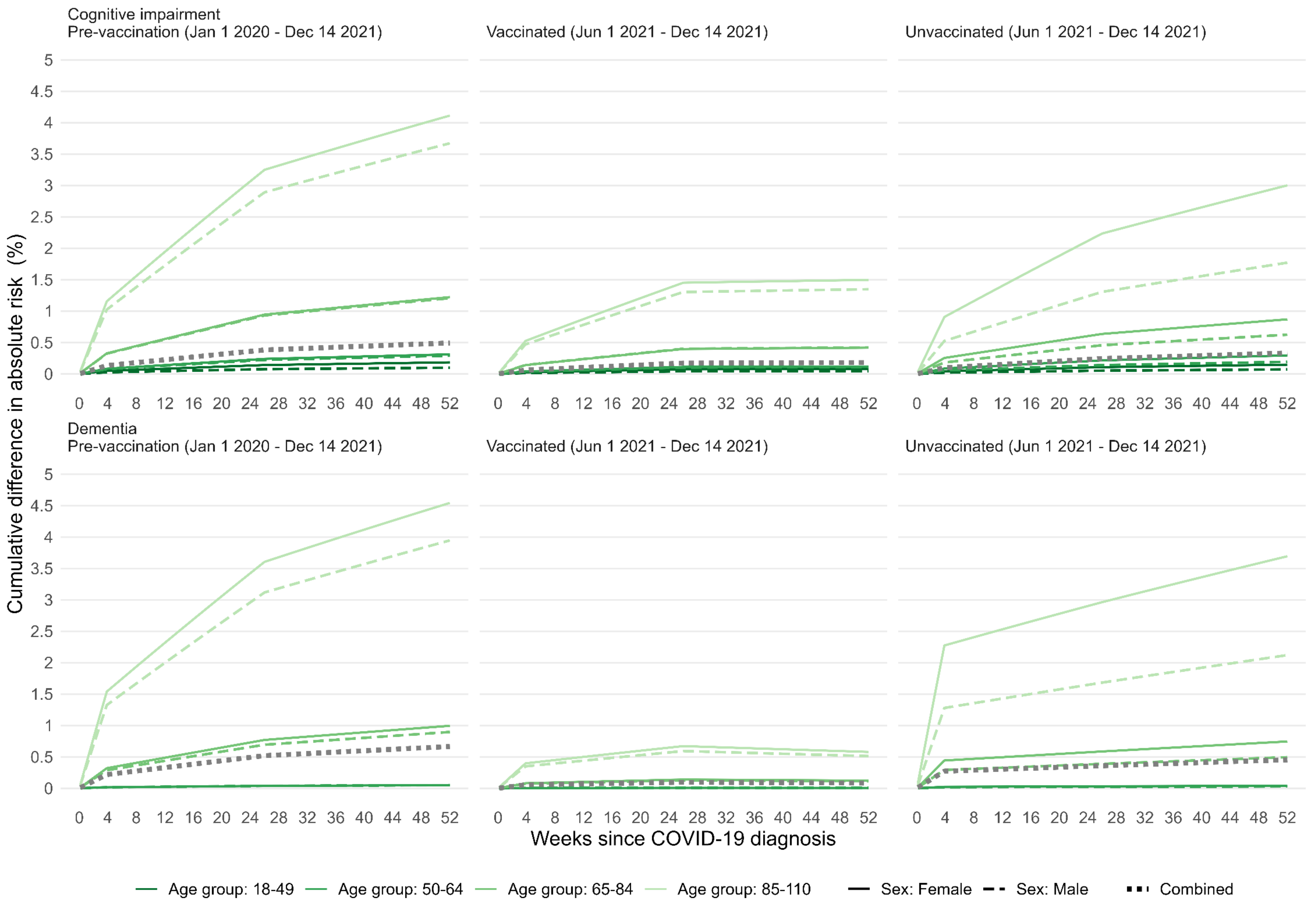
